# Severe fatigue as symptom of long COVID is characterized by increased expression of inflammatory genes in monocytes, increased serum pro-inflammatory cytokines, and increased CD8+ T-lymphocytes: A putative dysregulation of the immune-brain axis, the coagulation process, and auto-inflammation to explain the diversity of long COVID symptoms

**DOI:** 10.1101/2022.09.15.22279970

**Authors:** Julia C. Berentschot, Hemmo A. Drexhage, Daniel G. Aynekulu Mersha, Annemarie J.M. Wijkhuijs, Corine H. GeurtsvanKessel, Marion P.G. Koopmans, Jolanda Voermans, Majanka H. Heijenbrok-Kal, L. Martine Bek, Gerard M. Ribbers, Rita J.G. van den Berg-Emons, Joachim G.J.V Aerts, Willem A. Dik, Merel E. Hellemons

## Abstract

**Background:** A significant proportion of patients with SARS-CoV-2 infection develops long COVID with fatigue as one of the most disabling symptoms. We performed clinical and immunologic profiling of fatigued and non-fatigued long COVID patients and age and gender matched healthy controls (HCs).

**Methods:** We included 37 long COVID patients with and 36 without severe fatigue and assessed inflammation-related monocyte gene expression, serum levels of inflammatory cytokines, and leukocyte and lymphocyte subsets 3-6 months after hospital discharge, and followed clinical symptoms up to one year.

**Results:** Long COVID with fatigue represented a severe variant with many symptoms (median 9 [IQR 5.0-10.0] symptoms) and signs of cognitive failure (41%) and depression (>24%). Symptoms persisted up to one year follow-up. Fatigued patients showed increased expression of inflammatory genes in monocytes, increased serum IL-6, TNF-α, galectin-9, and CXCL10, and increased CD8+ T-lymphocytes compared to HCs.

Non-fatigued long COVID patients were arbitrarily divided in those with moderately severe disease (4 [2.5-5.0] symptoms, primarily impaired fitness, n=25) and those with mild disease (1 [1.0-2.0] symptom, n=11). Symptoms in non-fatigued long COVID patients persisted up to one year follow-up. Moderately severe patients showed reduced CD45RO^-^ naïve CD4^+^ T-lymphocytes and CD25+FOXP3+ regulatory CD4^+^ T-lymphocytes and limited monocyte and serum (galectin-9) inflammation. Mild patients showed monocyte and serum (IL-6, galectin-9) inflammation and decreased CD4^+^ T-lymphocyte subsets (T-helper 1 cells).

**Conclusion:** Long COVID with fatigue is associated with many concurrent and persistent symptoms up to one year after hospitalization and with clear signs of low grade inflammation and increased CD8^+^ T-lymphocytes. We showed that long COVID is a clinical and immunologic heterogeneous disorder. Diagnostic tools and personalized therapies combatting the diverse immune abnormalities might be required to alleviate the persisting disabling complaints of the patients.

## 1. Introduction

A significant proportion of patients develops long-lasting symptoms after acute corona virus disease 2019 (COVID-19). Different terms have been used to describe this condition, such as long COVID, post-acute COVID-19 syndrome, post-acute sequelae of COVID-19, long-haulers, or post COVID-19 condition.^1, 2^ In the current report we will use the term long COVID, consistent with most literature and long COVID is also the most commonly used terminology amongst patients.

Long COVID represents a wide spectrum of ⍰ often disabling ⍰ symptoms. Frequently reported symptoms are fatigue, impaired fitness, dyspnea, and cognitive impairment.^3-5^ Numerous studies showed the presence of these symptoms beyond 3 months after acute severe acute respiratory syndrome coronavirus 2 (SARS-CoV-2) infection, with evidence of persistence even two years after the acute illness.^6^ As patients with long COVID differ substantially regarding symptoms, severity, and recovery profile, attempts have been made to discern different clinical phenotypes of long COVID, without reaching consensus to date.^7-9^

Disabling fatigue is one of the most prominent and severe symptoms of long COVID. Among patients who had been hospitalized for acute COVID-19, proportions of up to 41% to 60% have been reported for patients that still suffer from fatigue one year after hospital discharge, without evident improvement beyond 6 months.^5, 10, 11^ Furthermore, the fatigue has been shown to negatively affect quality of life.^12^ This troubling problem thus requires in-depth evaluation regarding its pathogenesis, facilitating future interventions.

The prolonged fatigue state after acute COVID-19 shows clinical similarities with other post-infectious fatigue syndromes, such as that after Coxiella burnetii (Q fever) and Epstein-Barr virus (infectious mononucleosis) infection, and also shows similarities with myalgic encephalomyelitis/chronic fatigue syndrome (ME/CFS).^13-15^ The latter is characterized by severe fatigue lasting more than 6 months and involves debilitating symptoms such as post-exertional malaise and cognitive problems.^16^ Immune activation, such as increased levels of circulating cytokines and increases of CD8^+^ T-lymphocytes, has been found in both post-infectious fatigue conditions and ME/CFS and is thought to play a role in the pathophysiology of these conditions.^16, 17^ Given the clinical similarities with post-infectious fatigue syndromes and ME/CFS, a similar immune activation may play a pathogenic role in long COVID. To date, several studies have identified persistent inflammatory and T- and B-lymphocyte abnormalities among patients in the convalescent phase of COVID-19.^18-21^ However, a consistent and comprehensive association of these immune abnormalities with the diverse clinical symptoms of long COVID could not be established.^18-20, 22, 23^ Moreover, an in-depth immunological assessment among patients with and without the severe fatigue symptom of long COVID is lacking.

In the present study we examined patients who developed long COVID after hospitalization for COVID-19, and focused on the patients with the severe fatigue symptom. We performed an immunological assessment between 3-6 months after hospital discharge while clinical symptoms were longitudinally assessed up to one year post-discharge. We performed a comprehensive assessment of patient-reported outcome measures (PROMs). Immunologically we determined the expression of various sets of inflammation-related genes in circulating monocytes along with serum levels of inflammation-regulating cytokines and leukocyte and lymphocyte subsets related to inflammation regulation. These assays were chosen as these have previously been shown to be related to immunological changes in patients with various mental and somatic disorders.^24-30^ The outcomes were compared to long COVID patients without fatigue and to a group of matched healthy individuals.

## 2. Methods

### 2.1. Study participants and procedure

This cross-sectional study (IMMUNOFATIGUE) was carried out at Erasmus Medical Center (MC) Rotterdam, The Netherlands. The study was performed within a prospective cohort study (CO-FLOW) on long-term outcomes in adult patients who had been hospitalized for COVID-19 in The Netherlands.^31^ Patients were eligible to participate in the IMMUNOFATIGUE study if they visited the outpatient clinic at Erasmus MC between 3 and 6 months after hospital discharge for persistent COVID-19 sequelae. Demographics and clinical characteristics at hospital admission were retrospectively collected from electronic medical records.

A group of age and gender matched healthy controls (HCs) without a history of SARS-CoV-2 infection (self-reported) and without signs of chronic fatigue or depression was recruited among hospital visitors. All HCs had been vaccinated against COVID-19 at the time of collecting blood samples. All patients and HCs provided written informed consent before the start of study measurements. This study was approved by the Medical Ethics Committee of Erasmus MC (MEC-2020-1893).

### 2.2. PROMs

PROMs were collected in all patients as part of the CO-FLOW study at 3, 6, and 12 months post-hospital discharge.^27^ For the IMMUNOFATIGUE study the Beck’s Depression Inventory (BDI-21) was added. HCs were screened for fatigue and depression using the Fatigue Assessment Scale (FAS) and Hospital Anxiety and Depression Scale (HADS).

Patients filled out a symptom questionnaire (Corona Symptom Checklist [CSC], Supplementary Methods) to assess the presence of new or worsened symptoms following acute SARS-CoV-2 infection on a binary scale (yes or no). In this study we included a selection of 12 typical long COVID symptoms. Several symptoms (such as chest pain, sensory overload, and headache) could not be taken into account as they were added to this questionnaire in a later stage and contain incomplete data.^5^ Dyspnea was assessed with the Modified Medical Research Council (mMRC) Dyspnea Scale,^32^ the questionnaire scales the severity of dyspnea from 0 (no dyspnea) to 4 (severe dyspnea); scores ≥2 were considered representative for the presence of dyspnea symptom. The severity of fatigue was assessed using the FAS (score range 0-50) with a score ≥22 indicating substantial fatigue.^33^ Anxiety and depression were assessed with the HADS and BDI-21. The HADS consists of the subscales (range 0-21) anxiety and depression, a subscale score ≥11 is considered clinically significant.^34^ Depression was also assessed with the BDI-21 (score range 0-63), scores 0-13 denote minimal depression, 14-19 mild depression, 20-28 moderate depression, and 29-63 severe depression.^35, 36^ Post-traumatic stress disorder (PTSD) was measured with the Impact of Event Scale-Revised (IES-R),^37^ with a score (range 0-88) ≥25 representing clinically significant PTSD. Health-related quality of life (HR-QoL) was assessed with the 36-item Short Form survey (SF-36).^38^ The SF-36 measures general health status and consists of eight domains; each domain score ranges 0-100, with lower scores indicating more disability.

### 2.3. Laboratory methods

#### 2.3.1. Blood collection and preparation

Serum and sodium heparinized peripheral blood samples were collected from patients during clinical follow-up (March to October 2021) and from HCs between July and November 2021. Peripheral blood mononuclear cells (PBMC) were isolated by low-density gradient centrifugation using Ficoll-density separation within 8 hours after blood draw to avoid erythrophagy-related activation of the monocytes. Isolated PBMC were frozen in 10%-dimetylsulfoxide and stored in liquid nitrogen.

#### 2.3.2. Monocyte gene expression

Expression levels of monocyte genes were assessed using procedures that have been described in previous publications.^30^ Briefly, CD14^+^ monocytes were isolated from thawed PBMCs by magnetic cell sorting (Automacs Pro Miltenyi Biotec, Bergisch Gladbach, Germany) and RNA was isolated (Qiagen RNeasy mini kit). Subsequently, RNA (0,5 μg) was reverse transcribed (High Capacity cDNA Reverse Transcription Kit; Applied Biosystems, Thermo Fisher Scientific) to obtain cDNA for quantitative-polymerase chain reaction (q-PCR). qPCR was performed using TaqMan Gene expression assays (Applied Biosystems). The expression levels of genes were determined using the comparative cycle (CT) method. All values were normalized to the housekeeping gene ABL1 (ΔCT values). Gene expression values of patients were also expressed relative to the average ΔCT value of HCs (ΔΔCT values). Positive ΔΔCT values denote downregulation and negative ΔΔCT values denote upregulation of gene expression relatively to HCs. The following genes were evaluated: ABCA1, ABCG1, ADM, BAX, BCL10, BCL2A1, CCL2, CCL20, CCL7, CXCL2, DUSP2, EGR1, EGR2, EMP1, HMOX1, IFI44, IFI44L, IFIT1, IFIT3, IL1A, IL1B, IL1R1, IL6, MAFF, MAPK6, MRC1, MVK, MX1, MXD1, NR1H3, PTX3, SERPINB2, TNF, TNFAIP3.

#### 2.3.3. Cytokine and soluble cell surface molecule measurement

The following cytokines and soluble cell surface molecules were measured in serum: brain-derived neurotrophic factor (BDNF), C-C motif chemokine ligand (CCL)2 and C-X-C motif chemokine ligand (CXCL)9 and CXCL10, cluster of differentiation 163 (CD163), galectin-9, granulocyte macrophage-colony stimulating factor (GM-CSF), interferon (IFN)-α, IFN-β, IFN-γ, interleukin (IL)-6, IL-7, IL-10, IL-12, T-cell immunoglobulin and mucin domain 1 (TIM-1), and tumor necrosis factor-alpha (TNF-α). All, except for IL-6, were determined using a Luminex multiplex bead-based assay (R&D Systems Europe, Abingdon, United Kingdom). IL-6 levels were determined using high senstitive ELISA (apDia, Turnhout, Belgium). All assays were performed in accordance with the manufacturer’s protocol.

#### 2.3.4. Lymphocyte immunophenotyping

Absolute counts of total leukocytes (CD45^+^), Natural Killer (NK) cells (CD3^-^CD16+CD56^+^), B-lymphocytes (CD19^+^), T-lymphocytes (CD3^+^), CD8^+^ T-lymphocytes, and CD4^+^ T-lymphocytes were determined using a clinical laboratory ISO 15189 accredited method.

For the analysis of CD4^+^ T-lymphocyte subsets 1 × 10^6^ of defrosted PBMCs were stimulated for 4⍰h at 37⍰°C in RPMI-1640 culture medium with 50⍰ng/ml phorbol 12-myristrate 13-acetate (PMA; Sigma Aldrich, St. Louis, MO, USA) and 1.0⍰μg/ml ionomycin (Sigma) in the presence of Golgistop (BD Biosciences). A 8-color (membrane and intracellular) staining was performed to determine the percentages of CD4^+^ T lymphocyte subsets relative to total lymphocytes. CD4^+^ T-lymphocyte subsets were identified by their secreting cytokines: T helper (Th)1 (CD3^+^CD4^+^IFNγ^+^), Th2 (CD3^+^CD4^+^IL4^+^), Th17 (CD3^+^CD4^+^IL17A^+^). Regulatory T-lymphocytes (T_reg_) were identified by their transcription factor FOXP3 (CD3^+^CD4^+^CD25hiFOXP3^+^). We also measured the proportions of naïve (CD3^+^CD4^+^CD45RO^-^) and memory (CD3^+^CD4^+^CD45RO^+^) CD4^+^ T-lymphocytes; the former was indicated by subtracting the proportion of memory CD4^+^ T-lymphocytes from the total CD4^+^ T-lymphocytes.^39, 40^ All flowcytometric analyses were performed using a FACS Canto II instrument (BD Biosciences).

#### 2.3.5. Epstein–Barr virus (EBV) and cytomegalovirus (CMV) assessment

Active EBV and CMV were assessed in randomly selected patients from the subgroups of long COVID to assess whether symptoms could be attributed to viral reactivation, as suggested in literature.^41^ EBV and CMV DNA load was measured using internal controlled quantitative real-time Taqman PCR based on assays performed as published previously.^42, 43^ For EBV a value over the lower limit of detection of >100 IU/mL indicated presence of active virus, and for CMV this was >50 IU/mL.

### 2.4. Statistical analysis

Continuous variables are presented as median and interquartile range (IQR) and categorical variables as a number and percentage. Differences in demographic and clinical characteristics between groups were analyzed using a Kruskal-Wallis test for continuous variables and a Fisher-Freeman-Halton exact test for categorical variables. The number of clinical symptoms was calculated using 14 typical long COVID symptoms, 12 symptoms from the CSC and the symptoms fatigue (FAS) and dyspnea (mMRC Dyspnea Scale). Hierarchical cluster analysis of monocyte gene expression levels was performed using Spearman’s rank correlation coefficient matrix to identify clusters of mutually correlating genes. Missing gene values (0.7% of COVID-19 patients, 1.4% of HCs) were imputed with the median of patient or HC value. We used ΔΔCT values for single gene expressions, p-values were calculated with Wilcoxon signed rank test using the Benjamini-Hochberg-method for multiple testing. In the serum analysis, the majority of samples had GM-CSF, IFN-α, IL-10, IL-12, and TIM-1 levels below the lower limit of detection (LOD) (Supplementary Figure S1) and therefore these cytokines were excluded from further analysis. For other markers, values below the LOD were imputed by half of the lowest value observed of a given cytokine, that is in long COVID patients for IFN-γ (70% <LOD), IFN-β (64%), CXCL9 (39%), IL-6 (35%), IL-7 (19%). Extreme outliers (TNF-α n=1, IFN-β n=1, CXCL9 n=1) were removed due to potential erratic measurements. We performed a Kruskal-Wallis test and post-hoc test with Bonferroni correction for multiple comparisons to assess differences between long COVID variants and HCs in the serum cytokine and soluble cell surface molecule levels and leukocyte and lymphocyte subsets. We assessed the correlation between fatigue (FAS score) and CD8^+^ T-lymphocyte counts using Spearman’s rank correlation coefficient. We performed an additional data analysis on unsupervised hierarchical clustering using the most distinctive immune characteristics (monocyte genes IL-6, CCL20, CXCL2, CD8^+^ T-lymphocytes, and naïve CD4^+^ T-lymphocytes) to assess subgroups of patients on the basis of their immune profile, all values were expressed relative to the average of HCs (see Supplementary Results). A p-value <0.05 was considered statistically significant. Statistical analyses were performed with IBM SPSS Statistics version 28 (SPSS Inc., Chicago, IL, USA) and R software version 1.4.

## 3. Results

### 3.1. Participants

We included 37 long COVID patients with severe fatigue at the time of collecting blood samples. As a contrast group we included 36 long COVID patients who did not experience severe fatigue, yet showed other signs and symptoms of long COVID. All patients had been discharged from hospital between October 2020 and May 2021, representing patients with SARS-CoV-2 alpha variant. The median follow-up time (day of blood sampling) since hospital discharge was 107.0 (92.5-138.5) days. The patients’ demographic and clinical characteristics are shown in Supplementary Table S1 for the long COVID patients with and without severe fatigue. Among the fatigued long COVID patients, the median age was 59.0 (IQR 56.0-66.5) years, 24 (64.9%) were male, 18 (48.6%) had been treated in the intensive care unit (ICU) for acute COVID-19, and the length of stay (LOS) in hospital was 17.0 (9.0-26.0) days. Demographic and clinical characteristics did not differ between long COVID patients with and without fatigue (Supplementary Table S1). As controls we included 42 age and gender matched healthy controls (HCs) with a median age of 62.0 (51.8-68.3) years (p=0.9, comparison with long COVID fatigue group) and 26 (61.9%) were male (p=0.8).

### 3.2. Clinical characteristics of long COVID with fatigue

The PROMs in the fatigued long COVID variant are presented in Table 1. The patients experienced low energy levels throughout the day. Regarding the main symptoms, collectively, all patients reported ≥3 symptoms, including impaired fitness. Other PROMs showed that 40.5% of the patients experienced cognitive failure. Depression was found via HADS-D score in 24.3% and via BDI-21 score in 37.2% of the patients. This severe fatigue variant of long COVID demonstrated significantly worse symptomatology as compared to long COVID patients without fatigue and was further characterized by significantly lower HRQoL outcomes (Table 1). Figure 1 presents the recovery profile of the main symptoms during the first year after hospital discharge. The severe fatigue variant showed hardly any clinical improvement over time, as 95.0% of the patients reported ≥3 symptoms at one year follow-up.

**Table 1.**
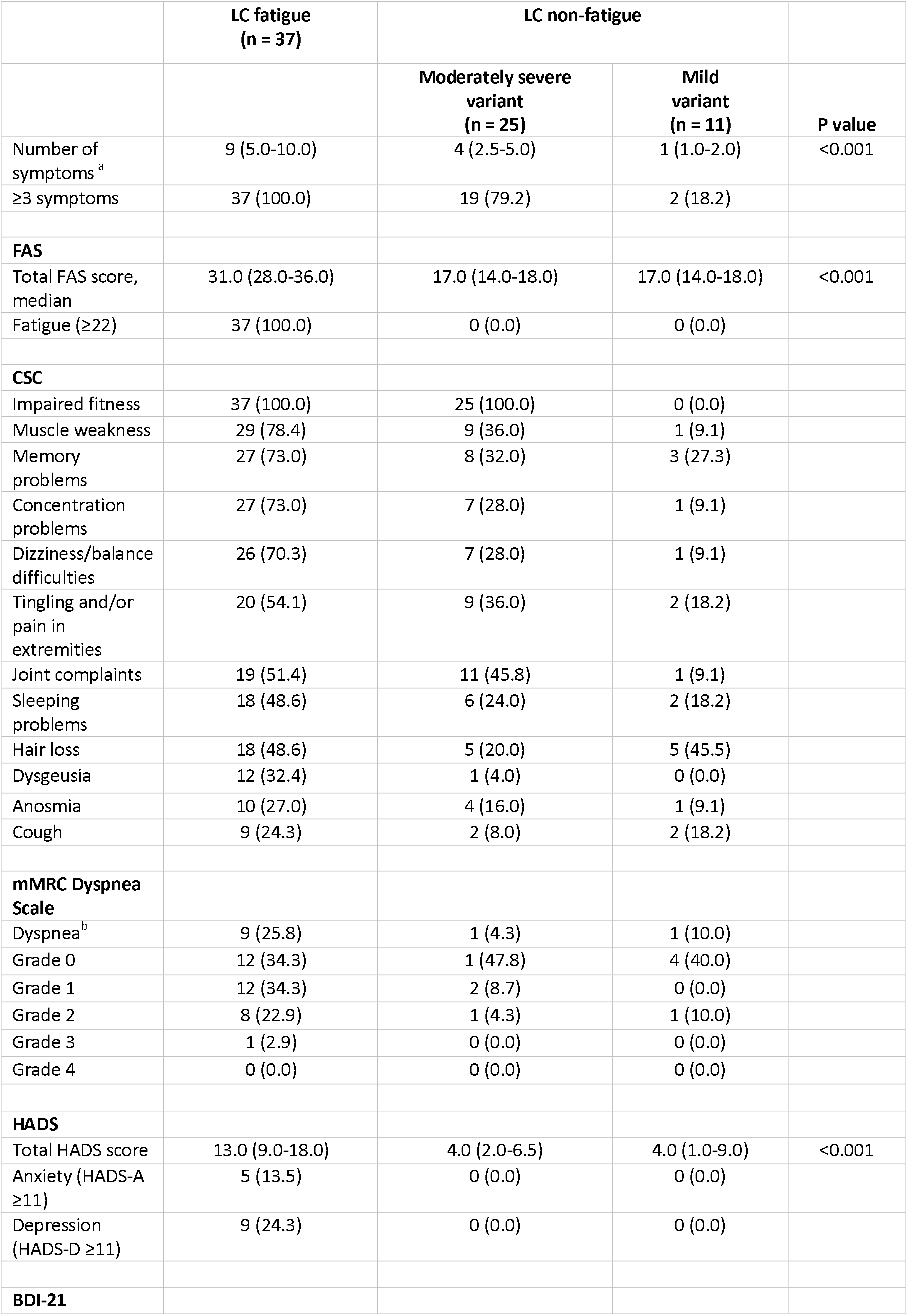
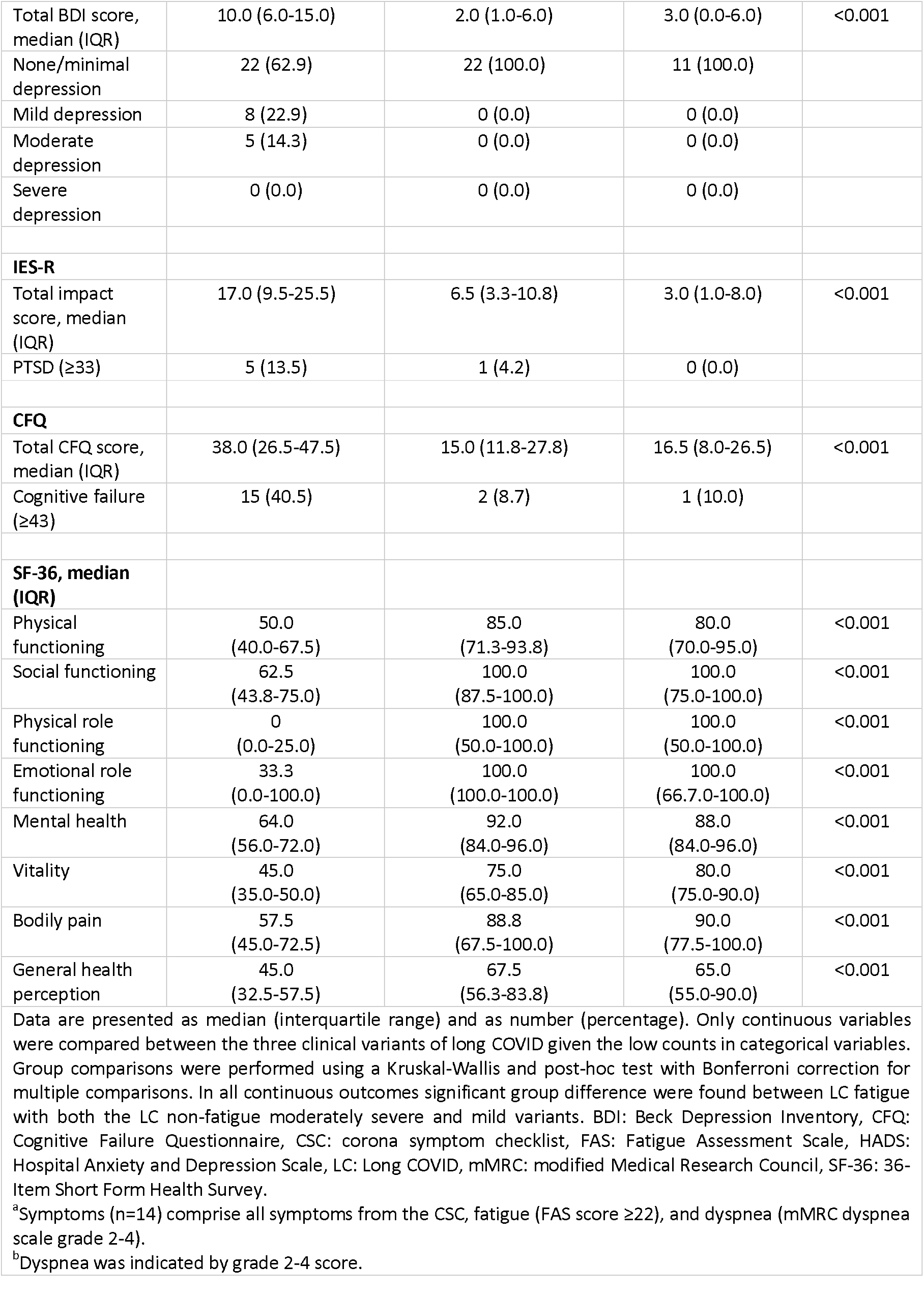
Patient-reported outcome measures.

**Figure 1.**
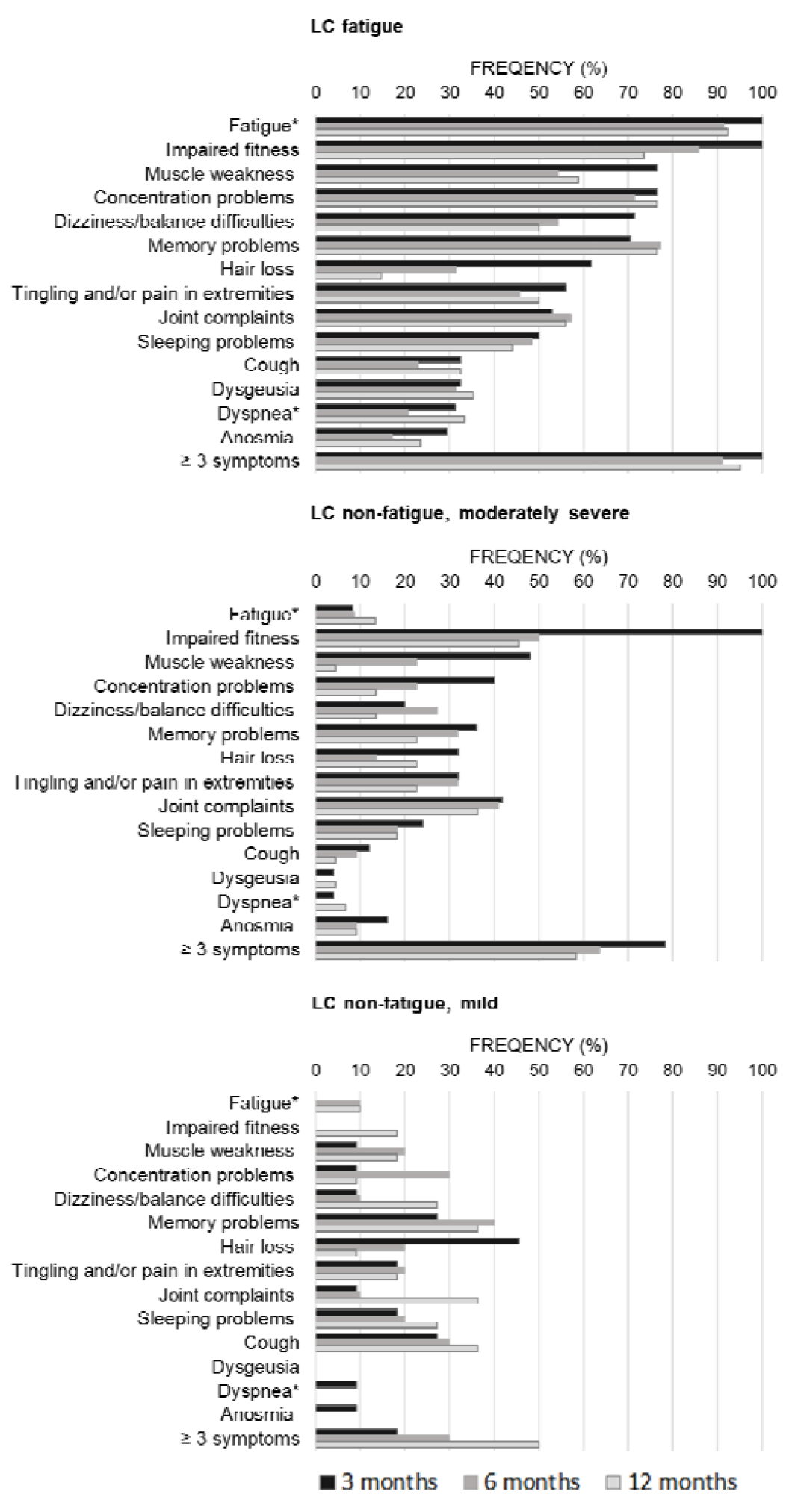
Prevalence of clinical symptoms across the three clinical variants of long COVID assessed at 3, 6, and 12 months after hospital discharge. Symptoms were obtained from the Corona Symptom Checklist on the presence of new or worsened symptoms following a SARS-CoV-2 infection (yes or no). * The fatigue symptom was obtained from the fatigue assessment scale questionnaire (total score >22 denotes fatigue)^29^ and the dyspnea symptom was obtained from the modified medical research council dyspnea scale (grade ≥2 denotes dyspnea)^28^. LC fatigue: patients experienced fatigue at the time of collecting blood samples. LC non-fatigue, moderately severe variant: patients experienced impaired fitness without fatigue. LC non-fatigue, mild variant: patients without impaired fitness and fatigue but reporting other symptoms. LC: Long COVID.

### 3.3. Clinical characteristics of long COVID without fatigue

Among the 36 long COVID patients without severe fatigue, impaired fitness was the most frequent symptom (n=25/36, 69.4%), and this symptom appeared indicative of severity and immunologic differences. We therefore arbitrary divided the long COVID patients without fatigue in two groups: a *moderately severe variant* of long COVID that consisted of patients with impaired fitness but without fatigue (25/73, 34.2% of all long COVID patients) and a *mild variant* that consisted of patients (11/73, 15.1%) with neither fatigue or impaired fitness. The PROMs in the moderately severe and mild disease variant are presented in Table 1.

#### Moderately severe variant

Patients with this variant experienced moderate-to-high energy levels throughout the day. Nevertheless the majority of patients (79.2%) reported ≥3 symptoms upon inclusion, reducing slightly to 58.3% at one year follow-up (Figure 1).

#### Mild variant

Patients with this variant experienced high energy levels throughout the day and a minority of the patients (18.2%) reported ≥3 symptoms, but this proportion increased to 50.0% during follow-up. One patient developed severe fatigue and two patients impaired fitness at one year follow-up.

### 3.4. Immune characteristics of long COVID with fatigue

### 3.4.1. Monocyte gene activation

Hierarchical clustering of monocyte gene expression levels (66 long COVID patients with complete data) revealed three main gene clusters (Figure 2A). These clusters represent strong mutually correlating genes within each cluster; only cluster C genes correlated weaker amongst themselves. Gene cluster A was composed of inflammation-regulation genes and genes related to adhesion, chemotaxis, apoptosis, and pyroptotic mechanisms of the cells. Cluster B consisted of type 1 IFN driven inflammation-related genes (a cluster we found upregulated in systemic autoimmune conditions).^44, 45^ Cluster C consisted of genes involved in mitochondrial anti-inflammatory action and cholesterol pump genes (a hall mark cluster of monocytes in patients with major depressive disorder [MDD]).^30^ The found clusters are similar to the upregulated gene clusters found in previous studies in MDD, bipolar disorder (BD), and various autoimmune disorders (thyroid autoimmune disease, type 1 diabetes, Sjögren disease and SLE).^20-30^

**Figure 2.**
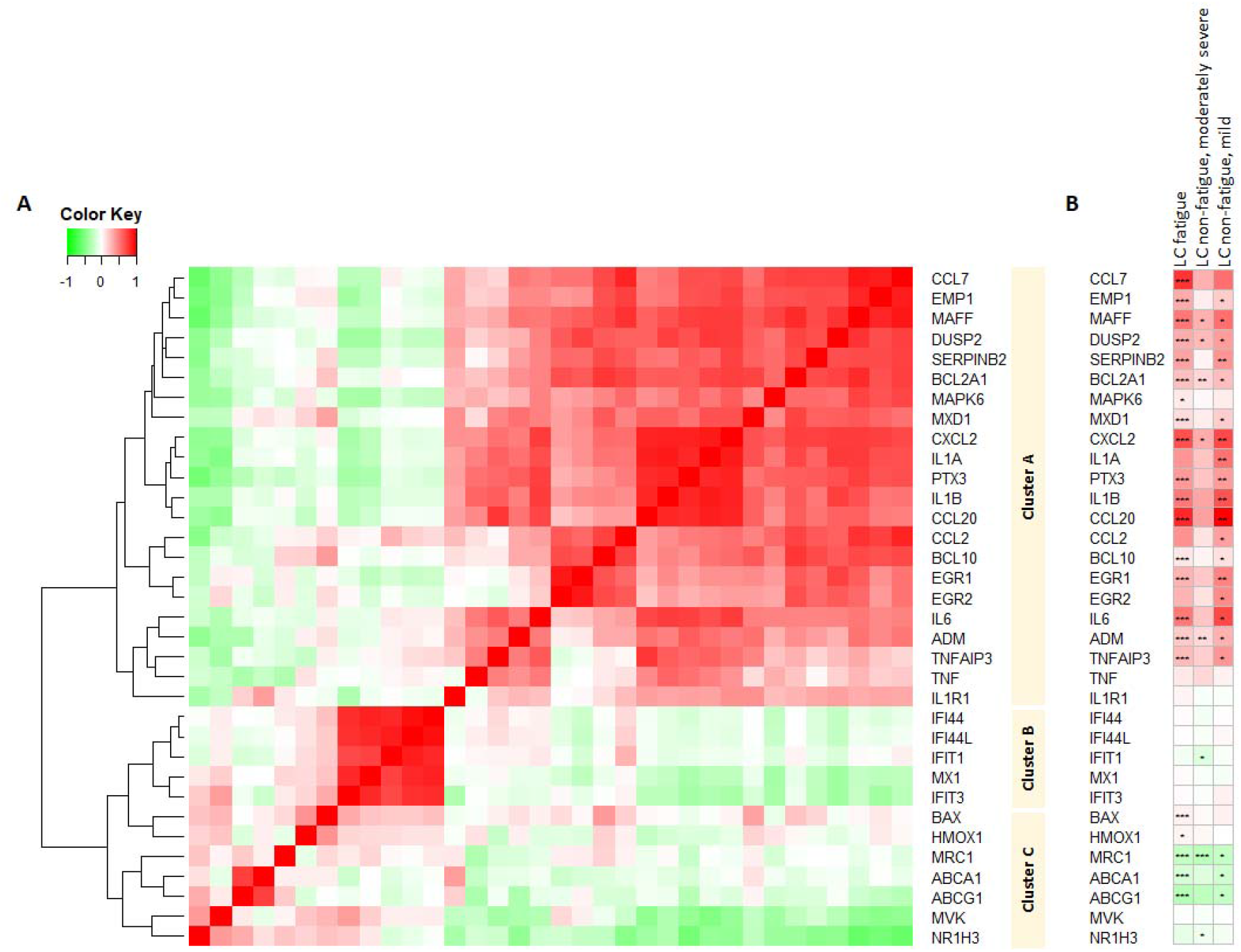
Hierarchical clustering and heatmap of the expression of monocyte genes in long COVID patients (2A) and single gene expressions across three variants of long COVID (2B) 2A: Three main gene clusters can be identified. Cluster A comprises inflammation-regulation genes and genes related to adhesion, chemotaxis, apoptosis, and pyroptotic mechanisms of the cells. Cluster B comprises genes related to type 1 interferon driven inflammation. Cluster C comprises mainly genes related to mitochondrial anti-inflammatory action and cholesterol pump genes. Correlations between genes were measured using Spearman rank correlations. 2B: Single gene expression levels across the three variants are presented as mean values and are expressed relatively to the expression level of healthy controls; the intensity of red reflects higher expression (upregulation) and green reflects lower expression (downregulation). P-values were calculated with Wilcoxon signed rank test using the Benjamini-Hochberg-method for multiple testing, the level of significance is indicated by *p<0.05, **p<0.01, ***p<0.001. LC fatigue: patients experienced fatigue at the time of collecting blood samples. LC non-fatigue, moderately severe variant: patients experienced impaired fitness without fatigue. LC non-fatigue, mild variant: patients without impaired fitness and fatigue but reporting other symptoms. LC: Long COVID.

Figure 2B shows the monocyte gene expression pattern in the severe fatigue variant relatively to the expression levels of HCs. Patients with this variant were characterized by many significantly overexpressed cluster A inflammation-regulating genes (e.g. IL-1B, IL-6, CCL7, CCL20) as well as some cluster C genes (BAX, HMOX1) (Figure 2B). The cholesterol pump genes (ABCA1, ABCG1) and the M2 marker MRC1 were significantly downregulated. Normal expression levels were found for the type 1 IFN induced genes (ISGs) in cluster B. This profile represents a strong pro-inflammatory state of the monocytes.

#### 3.4.2. Serum cytokine and soluble cell surface molecule levels

The level of the various tested inflammatory cytokines was evaluated to further investigate the inflammatory state of the long COVID patients with fatigue (Supplementary Figure S1 and Figure 3). Figure 3 presents only the outcomes for the cytokine and soluble cell surface molecule levels we found to differ from those of HCs.

**Figure 3.**
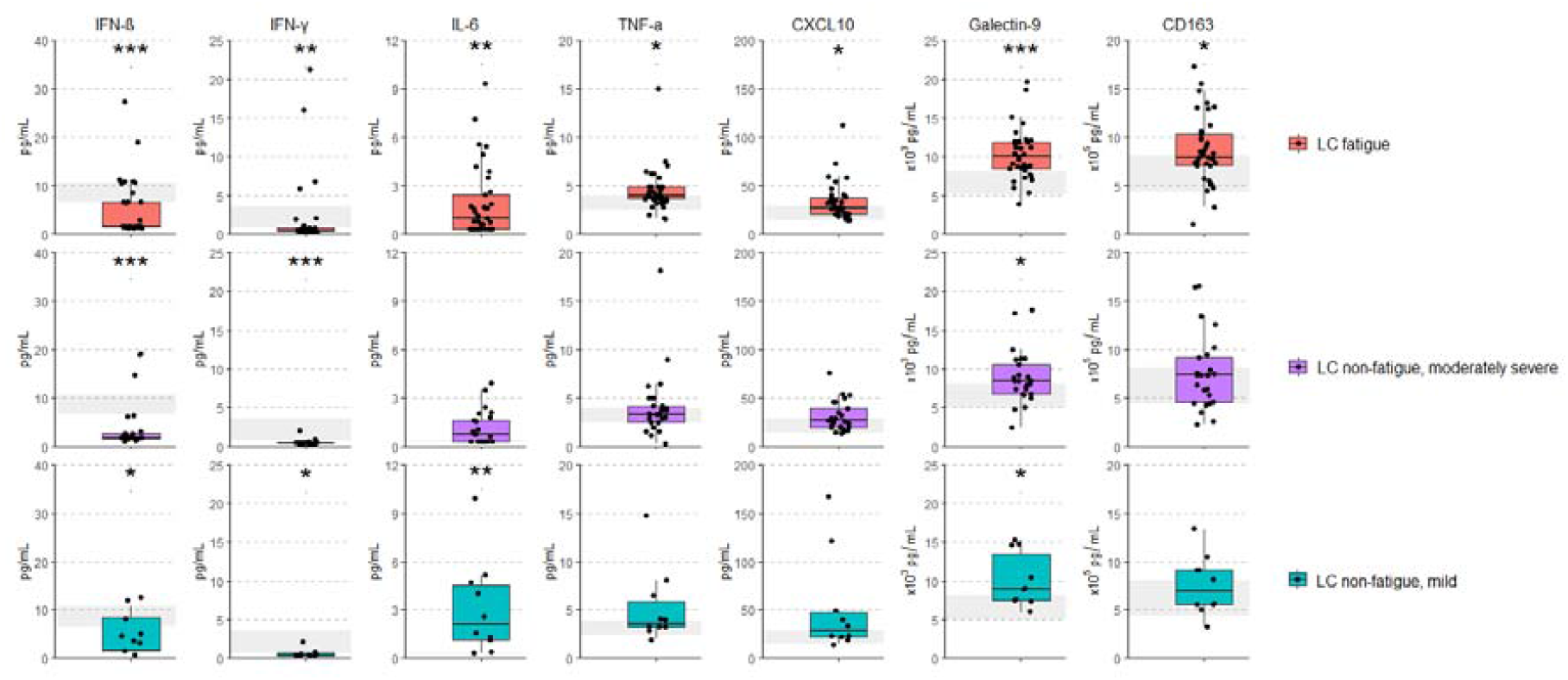
Significant group differences in serum cytokine and soluble cell surface molecule levels between the three clinical variants of long COVID with healthy controls. Group comparisons in serum cytokine and soluble cell surface molecule levels between long COVID variants and healthy controls were performed with the Kruskal-Wallis test and post-hoc test with Bonferroni correction for multiple comparisons. The grey coloured area represents the interquartile range of the measured cytokine level in healthy controls. A significant group difference between each variant with the healthy control group is presented, denoted by *p<0.05, ** p<0.01, ***p<0.001. LC fatigue: patients experienced fatigue at the time of collecting blood samples. LC non-fatigue, moderately severe variant: patients experienced impaired fitness without fatigue. LC non-fatigue, mild variant: patients without impaired fitness and fatigue but reporting other symptoms. LC: Long COVID. BDNF: brain-derived neurotrophic factor, CCL: C-C motif chemokine ligand, CXCL: C-X-C motif chemokine ligand, CD163: cluster of differentiation, IL: interleukin, GM-CSF: granulocyte macrophage-colony stimulating factor, IFN: interferon, TIM: T-cell immunoglobulin and mucin domain 1 (TIM-1), TNF-a: tumor necrosis factor-alpha.

Concerning IFN-driven inflammation, reduced IFN-β and IFN-γ levels were present in all long COVID groups compared to HCs (Figure 3). The severe fatigue variant was also characterized by significantly higher levels of IL-6, TNF-α, CXCL10, galectin-9, and CD163 as compared to HCs. These findings further support a strong pro-inflammatory state of monocytes in this variant.

#### 3.4.3. Circulating leukocyte and lymphocyte subsets

We performed enumerations of the number of circulating leukocytes, T- and B-lymphocytes, and NK cells. With regard to B-lymphocytes, NK cells, and CD4^+^ T-lymphocytes, significant differences between long COVID groups and HCs were not found (Supplementary Figure S2). In the severe fatigue variant the numbers of leukocytes and total T-lymphocytes were significantly raised as compared to HCs (Supplementary Figure S2), these increases were primarily due to an increase in CD8^+^ T-lymphocytes (Figure 4A). Higher numbers of CD8^+^ T-lymphocytes associated significantly with more severe fatigue among all long COVID patients, although the correlation coefficient is weak (Figure 4B).

**Figure 4.**
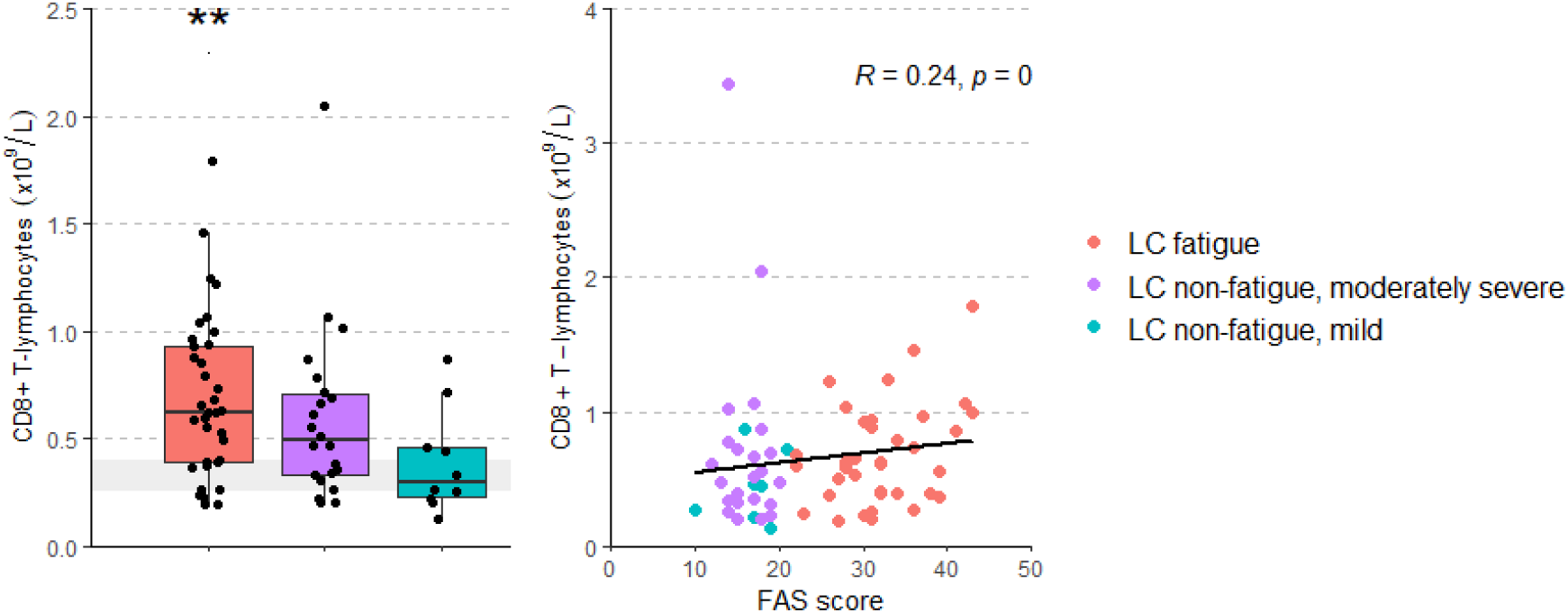
CD8^+^ T-lymphocyte numbers across the three clinical variants of long COVID compared with healthy controls (A) and the correlation between CD8^+^ T-lymphocytes with the level of fatigue (B) 4A: Group comparison between the three clinical variants of long COVID and healthy controls in CD8^+^ T-lymphocyte (CD3^+^CD8^+^) numbers was performed with the Kruskal-Wallis test and post-hoc test with Bonferroni correction for multiple comparisons. The grey coloured area represents the interquartile range of the measured CD8^+^ T-lymphocyte numbers in healthy controls. A significant group difference between each variant with the healthy controls is presented, **p<0.01. 4B: Spearman’s Rho correlation between the number of CD8^+^ T-lymphocytes and the level of fatigue assessed with de FAS; a higher FAS Score represents more severe fatigue. LC fatigue: patients experienced fatigue at the time of collecting blood samples. LC non-fatigue, moderately severe variant: patients experienced impaired fitness without fatigue. LC non-fatigue, mild variant: patients without impaired fitness and fatigue but reporting other symptoms. FAS: fatigue assessment scale, LC: Long COVID.

With regard to the percentages of CD4^+^ T-lymphocyte populations (Supplementary Figure S3), outcomes did not significantly differ between the severe fatigue variant and HCs. In contrast, differences in the CD4^+^ T-lymphocyte populations were found between long COVID patients without fatigue in comparison with HCs (Figure 5).

**Figure 5.**
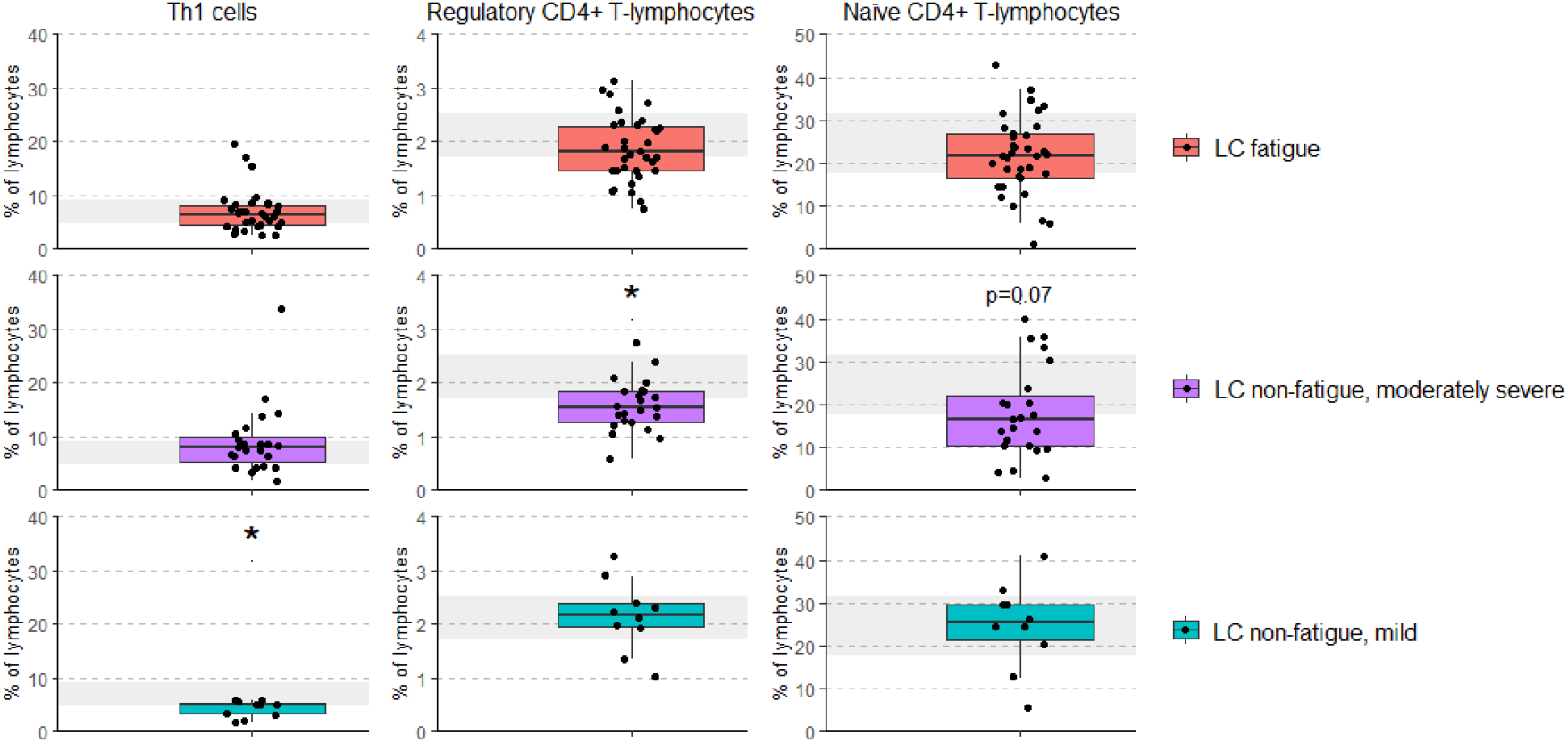
Significant group differences in CD4^+^ T-lymphocyte subsets between the three clinical variants of long COVID with healthy controls. Group comparisons between three variant of long COVID and healthy controls in the percentages of T helper 1 cells (CD3^+^CD4^+^IFNγ^+^), regulatory CD4^+^ T-lymphocytes (CD3^+^CD4^+^CD25hiFOXP3^+^), and naïve CD4^+^ T-lymphocytes (CD3^+^CD4^+^CD45RO^−^) were performed with the Kruskal-Wallis test and post-hoc test with Bonferroni correction for multiple comparisons. Significant differences between each variant with healthy controls are presented, a significant difference is denoted by *p<0.05. The grey coloured area represents the interquartile range of the measured outcome in healthy controls. LC fatigue: patients experienced fatigue at the time of collecting blood samples. LC non-fatigue, moderately severe variant: patients experienced impaired fitness without fatigue. LC non-fatigue, mild variant: patients without impaired fitness and fatigue but reporting other symptoms. LC: Long COVID.

### 3.5. Immune characteristics of long COVID without fatigue

#### 3.5.1. Monocyte gene activation

Figure 2B shows the monocyte gene expression pattern in the moderately severe and mild non-fatigue long-COVID, relatively to expression levels of HCs. Similar to the severe fatigue variant, normal expression levels were found for ISGs in cluster B.

##### Moderately severe variant

These patients showed some monocyte activation with only 5 cluster A genes significantly raised. Downregulated expression levels were found for only one ISG (IFIT1) in cluster B and for the cluster C genes MRC1 NR1H3 (Figure 2B).

##### Mild variant

This variant showed a similar pattern of monocyte inflammatory activation as the severe fatigue variant (Figure 2B), and thus represents a clear pro-inflammatory state of the monocytes.

#### 3.5.2. Serum cytokine and soluble cell surface molecule levels

Similar to the severe fatigue variant, reduced serum IFN-β and IFN-γ levels were present in the moderately severe and mild variants compared to HCs (Figure 3). Other levels that significantly differed from HCs in the moderately severe and mild variant are reported below.

##### Moderately severe variant

Only galectin-9 levels were significantly upregulated in this variant as compared to HCs, with near significant raised levels of IL-6, indicating a minor pro-inflammatory state as compared to the other variants.

##### Mild variant

The cytokine pattern appeared similar to the pattern of the severe fatigue variant, be it to a slightly lesser degree, of note: thus similar to the findings with the inflammatory gene expression. We found significantly higher levels of IL-6 and Galectin-9 compared to HCs in this variant.

### 3.5.3. Circulating leukocyte and lymphocyte subsets

The numbers of leukocytes, total T- and B-lymphocytes, NK cells, and CD8^+^ and CD4^+^ T-lymphocytes did not differ significantly from HC values in the moderately severe and mild variants (Supplementary Figure S2). Regarding the CD4^+^ T-lymphocyte populations, significant differences between the moderately severe and mild variants with HCs were not found in the percentages of Th2 cells, Th17 cells, and memory CD4^+^ T-lymphocytes (Supplementary Figure S3).

Figure 5 shows the percentages of Th1 cells, CD4^+^ T_reg_ lymphocytes, and naïve CD4^+^ T-lymphocytes, populations which were found to differ significantly between long COVID variants and HCs.

#### Moderately severe variant

These patients showed the strongest abnormalities in the CD4^+^ T-lymphocyte populations. The percentage of naïve CD4^+^ T-lymphocytes and CD4^+^ T_reg_ lymphocytes were downregulated as compared to HCs.

#### Mild variant

Percentages of Th1 cells were significantly downregulated in this variant compared to HCs as well as compared to the moderately severe variant (Supplementary Figure S3).

### 3.6. No signs of EBV and CMV reactivation in long COVID patients

Viral load of EBV and CMV were tested in randomly selected 10 (27%) patients of long COVID with fatigue, 10 (40%) of long COVID without fatigue, moderately severe variant, and 9 (82%) of long COVID without fatigue, mild variant. We did not find detectable viral loads in any of these patients, and thus did not perform further tests on the remaining patients.

### 3.7. Unsupervised hierarchical cluster analysis using immune characteristics in long COVID patients

Above we described the immune alterations of long COVID patients with and without fatigue at an average group level. Additionally we performed an unsupervised hierarchical cluster analysis to assess the patients on the basis of their immunotype. The analysis is further described in the Supplementary Results section. We were able to divide the patients into six different immunotypes (Figure S5A) and showed that trends of a preferred link of the clinical variants of long COVID to specific immunotypes were visible (Figure S5B and Table S2).

## 4. Discussion

This is the first study that focused on long COVID patients with severe fatigue, and provides insight into the clinical and immunologic profiles of long COVID patients with and without fatigue. Long COVID presenting with severe fatigue is clinically associated with many concurrent and persistent symptoms beyond one year and immunologically with clear signs of immune alterations, characterized by low grade inflammation and high levels of CD8^+^ T-lymphocytes. Long COVID patients without fatigue also showed immune alterations, such as minimal signs of low grade inflammation and altered CD4^+^ T-lymphocyte subsets. Long COVID appears to be a heterogeneous disorder regarding clinical symptoms and aberrant immune profiles, and despite several trends, we were unable to link specific long COVID symptoms to specific immune abnormalities.

### 4.1. Long COVID with fatigue

The long COVID patients with severe fatigue experienced many concurrent symptoms and showed a high burden of long COVID disease one year after hospital discharge. Signs of depression were only found in the severe fatigue variant of long COVID (24-37%). Immunologically long COVID with fatigue was characterized by an increased expression of inflammatory genes in monocytes, increases in serum inflammatory cytokine and soluble cell surface molecule levels, and an increase in CD8^+^ T-lymphocytes. Also Peluso and colleagues found higher levels of pro-inflammatory cytokines in the circulation 3 and 6 months after acute infection, in particular in those patients with the highest number of long COVID symptoms,^23^ while Phetsouphanh and colleagues reported that patients with long COVID had highly activated innate immune cells.^20^ In the unsupervised hierarchical clustering the majority of our severe patients indeed fell in the immunotypes characterized by increased expression of inflammatory genes in monocytes. Thus low grade inflammation was a definite sign of this severe fatigue variant of long COVID.

With regard to the increase in CD8^+^ T-lymphocytes, these cells play an important role in the host defense against viral infection by secreting cytokines and killing infected cells. Townsend and colleagues also found a long lasting increase in CD8^+^ T-lymphocytes in convalescent patients (up to 3 months) but could not find an association with persistent complaints of long COVID.^22^ Shuwa and colleagues however reported that T-lymphocytes displayed continued alterations with persistence of a cytotoxic program evident in CD8^+^ T-lymphocytes in patients with poor clinical outcomes at 6 months follow-up.^18^ Unfortunately we did not perform an in-depth analysis of the CD8^+^ T-lymphocyte subsets to further characterize the type of CD8^+^ T-lymphocytes which was increased in our patients. This may have guided us more into the cause of the increase. It is tempting to speculate that the increased numbers of CD8^+^ T-lymphocytes in the most severe clinical variant represent an ongoing “hidden” SARS-CoV-2 infection.

Apart from ongoing SARS-CoV-2 infection, it is also possible that other chronic infections might be involved in this type of immune activation, since other studies on long COVID have shown reactivation of herpesviruses.^41^ As we were unable to find signs of EBV and CMV reactivity in the patient, we favor the idea that the increased CD8^+^ T-lymphocyte numbers may indeed indicate persistent SARS-CoV-2 infection, in line with current hypotheses on viral persistence as a potential causal factor in long COVID.^46^

### 4.2. Long COVID without fatigue

The long COVID patients who did not suffer from severe fatigue at the time of collecting blood samples reported considerable other symptoms. With regard to the moderately severe variant, clinically this variant showed some improvement in the period of follow-up and the impaired fitness disappeared in 60% of the patients one year after hospital discharge. Still, more than half of the patients reported ≥3 symptoms at one year follow-up. Immunologically the moderately severe variant was characterized by a borderline significant decrease in the percentage of CD45RO^-^ naïve CD4^+^ T-lymphocytes (hereafter called a naïve CD4^+^ lymphocytopenia) and reduced CD4^+^ T_reg_ lymphocytes. Phetsouphanh and colleagues also found that a proportion of their long COVID patients lacked naïve T-lymphocytes (and also naïve B-lymphocytes).^20^ In our unsupervised hierarchical clustering the immunotypes characterized by a naïve CD4^+^ lymphocytopenia indeed harbored the majority of moderately severe patients. The moderately severe variant only showed limited monocyte inflammatory gene activation, while serum galectin-9 was significantly increased and IL-6 nearly significantly increased. Thus, signs of low grade inflammation were present, but not outspoken, and CD4^+^ T-lymphocyte subset abnormalities were clearly more prominent.

A reduction of naïve CD4^+^ T-lymphocytes is often considered as one of the signs of immunosenescence, the aging of the immune system.^47^ Also Wiech and colleagues reported in convalescent patients 6 months after severe COVID disease an immunosenescent profile of particularly the CD8^+^ T-lymphocyte population, but the authors were unable to find an association of this abnormality with long lasting complaints.^19^ An early premature aging of the immune system is known to be induced by chronic viral infections; chronic CMV infection is a well-known example of such aging.^19, 48^ It is thus tempting to speculate that also the CD4^+^ T-lymphocyte abnormalities of this moderately severe clinical variant are caused by ongoing SARS-CoV-2 infection, while also inducing some signs of limited low grade inflammation.

With regard to the clinically mild variant of long COVID without fatigue, although symptoms were overall less prevalent patients did in general not improve, 5/11 patients worsened while only two patients completely recovered at one year follow-up. Surprisingly all of these patients showed clear signs of low grade inflammation: Monocyte inflammatory gene activation and serum levels of IL-6 and Galectin-9 were raised in virtually similar way as in the severe fatigue variant of long COVID. In contrast to the severe fatigue variant CD8^+^ T-lymphocytes were not increased, while Th1 cells were decreased. In the unsupervised hierarchical clustering we found that the majority of mild patients were in immunotypes lacking naïve CD4^+^ T-lymphocytopenia and one of these immunotypes even showed CD4^+^ *T-lymphocytosis* with a prevalence of 40% of the mild patients. It is thus tempting to hypothesize that this relatively mild clinical form is an immunologically milder form of the severe fatigue form with pro-inflammatory mechanisms active, but not aggravated by the CD8^+^ T-lymphocytes and premature CD4^+^ T-lymphocyte senescence, and kept in balance by anti-inflammatory phenomena (Th1 cell reductions).

### 4.3. Comparison of the immune profile of long COVID fatigue to that of Myalgic Encephalomyelitis/Chronic Fatigue Syndrome (ME/CFS) and major depressive disorder (MDD). A disturbed immune-brain axis?

The fatigue of long COVID is reminiscent of the fatigue of ME/CFS. In our study fatigue associated primarily with monocyte inflammatory gene activation, high numbers of CD8^+^ T-lymphocytes, and increased serum levels of pro-inflammatory cytokines. Similar immune abnormalities have also been reported in ME/CFS patients.^16^

Fatigue is also an important component of MDD and it has been shown that SARS-CoV-2 infection is able to elicit episodes of major depression.^49, 50^ Indeed, also in our series we found a prevalence of 23% (HADS-D) and 37% (BDI-12) of depression in the group of long COVID patients with disabling fatigue. Our long COVID patients show monocyte gene expression abnormalities similar as those found in “regular” MDD patients with severe and recurrent depression.^30^

Also an early aging of the CD4^+^ T-lymphocyte compartment, similar to that found in the moderately severe impaired fitness form, occurs in MDD.^51,25^ Likewise, high CD8^+^ T-lymphocytes counts play a role in MDD.^52, 53^ The similarity of immune abnormalities between long COVID fatigue, ME/CFS, and MDD are thus striking. Considering these similarities it is tempting to speculate that similar immunopathogenic mechanisms may play a role in these disorders.

In MDD it is thought that the inflammatory activation of the circulating monocytes represent an activation of the myeloid system in general, including the microglia in the brain,^54^ particularly in areas regulating emotion.^55^ Increased pro-inflammatory cytokines in the serum of MDD patients induce pro-inflammatory IL-6 cascades in the brain, hypothesized to further activate the microglia, compromising their function.^56 57^ Also a normal functioning T-lymphocyte system is essential to regulate mood: Mice without functioning T-lymphocyte system show signs of anxiety and depressive-like behavior.^58^ Last, inflammatory cytokines and CD4^+^ T-lymphocyte subsets are correlated with structural brain changes in MDD.^55, 56^

With regard to ME/CFS there are also indications of a microglial inflammatory activation in the brain stem affecting the neuronal circuits regulating energy and metabolism. Brain stem scans showed structural and functional abnormalities^59^ that are very much alike to abnormalities found in long COVID patients.^60-62^

In our opinion further investigations should focus on a correlation of COVID-19 related structural and functional brain abnormalities, microglial activation, and the here described monocyte, T-lymphocyte, and serum inflammatory abnormalities to clarify a putative inflammation-induced abnormality in the central regulation of energy metabolism in the brain stem and mood regulation in the limbic system of fatigued long COVID patients.

### 4.4. Other putative pathogenic mechanisms in long COVID driven by the low grade inflammatory state

Apart from pathogenic mechanisms involving the brain, pathogenic mechanisms involving somatic processes have also been proposed as the basis for the symptoms of long COVID. Endotheliopathy^63^ and microvascular thrombosis^64^ are amongst these mechanisms. It is known that endothelial damage and thrombus formation does occur in the interaction of inflammatory monocytes with endothelial cells. Interestingly, the inflammatory monocyte gene expression signature was found raised in both the severe fatigued and relatively mild variant of long COVID and the signature contains molecules that play a role in the interaction of monocytes with endothelial cells, such as the adhesion EMP-1^65^ and the coagulation regulator SERPINB2, encoding PAI-2.^66^ Moreover, the pro-inflammatory cytokines induce processes perturbing the coagulation process.^67^ We therefore favor the idea that the here reported state of low grade monocyte inflammation may also impact vascular integrity and thrombus formation in long COVID patients and this process might be responsible for or contribute to symptoms, such as pains syndromes and muscle weakness.

Furthermore, the monocyte inflammatory state, the high serum cytokines, and the CD4^+^ T_reg_ lymphocyte deficiency might result in auto-inflammatory phenomena. A recent review listed a number of reports of inflammatory arthritis after SARS-CoV-2 infection, and also showed vasculitis and myositis to occur after COVID.^68^ In analogy to ME/CFS putative auto-antibodies to adrenergic, muscarinic, and G-protein coupled receptors might destabilize the autonomous nervous system leading to POTS-like (postural orthostatic tachycardia syndrome) and microvascular symptoms.^69^ Our group has extensively reported on a pro-inflammatory state of the myeloid cell system together with defects in the Treg lymphocyte system (as described here) as causal to the development of autoimmune thyroid disease, autoimmune diabetes, and autoimmune sialo-adenitis both in animal models as well as in humans.^25, 70-74^ Figure 6 sums up the here discussed hypothetical pathogenesis of the various signs and symptoms of long COVID caused by the here found monocyte and T-lymphocyte abnormalities.

**Figure 6.**
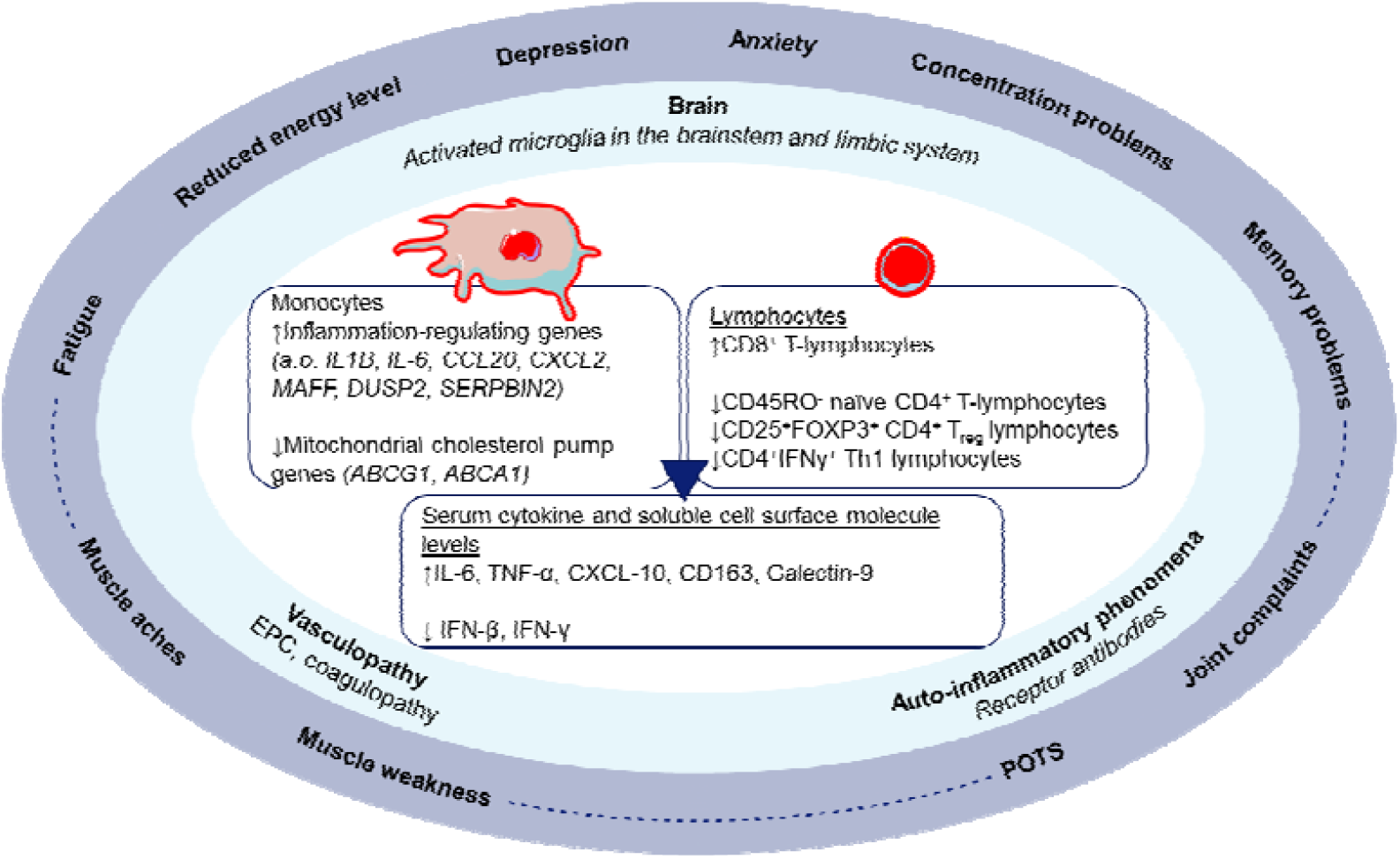
Hypothetical model of the pathogenesis of the diverse clinical signs and symptoms of long COVID. In the center are the here described pro-inflammatory monocyte, T-lymphocyte, and serum cytokine and soluble cell surface molecule abnormalities. In major depression and bipolar disorder these peripheral immune abnormalities correlate to inflammatory brain stem, limbic system and fronto-limbic system abnormalities, while in type 2 diabetes similar immune abnormalities resulted in the generation of reduced numbers of endothelial precursor cells and coagulapathies, leading to vasculopathies. Our group has also extensively published on similar pro-inflammatory monocyte/macrophage and T-lymphocyte dysregulations as causal to the generation of autoimmune reactions in humans with and in animal models of thyroid, islet and sialo-adenitis (see text for references). EPS: Endothelial progenitor cells; POTS: Postural tachycardia syndrome.

### 4.5. Therapeutic implications

The long term disabling consequences of long COVID with and without fatigue make a search for adequate treatment an urgent need. We postulate that the immune abnormalities found in this study form immune derangements which could be targeted in individualized treatments to combat the long term sequelae of SARS-CoV-2 infection. Although these immune abnormalities have not been proven as causal for the long COVID symptoms, lessening the symptoms by correcting the immune abnormalities would form indirect proof of causality. Interventions should be Individualized since patients show a diversity of immune abnormalities divided over the described immunotypes. Anti-inflammatory agents, such as COX-2 inhibitors, minocycline, dexamethasone or even anti-IL6, might be instrumental in dampening the excessive inflammatory processes. Effects could be monitored by testing the serum levels of galectin-9 and IL-6 and the monocyte inflammatory gene expression.

Agents stimulating type 1 IFN production, such as TLR-7 and TLR-9 stimulators (e.g. rintatolimod) might be instrumental in inducing IFN production and effects could be monitored by testing the ISG expression in monocytes and the serum level of IFN-β. Rintatolimod has been used with some success in ME/CFS.^75, 76^

Low dose IL-2 might be instrumental in correcting the reduced CD4^+^ T_reg_ lymphocytes and reduce naïve CD4^+^ T-lymphocytes. Low-dose IL-2 is a well-tolerated intervention and has proved to be instrumental in such corrections, particularly in systemic autoimmune states.^77, 78^

Above described immune correcting agents may have to be combined with an antiviral agent such as nirmatrelvir/ritonavir to combat the putative hidden viral reservoirs. Studies should be undertaken to further confirm the role of this putative reservoirs. One could argue that studies to test these putative treatments should not be further delayed to limit the devastating sequelae of long COVID.

### 4.6. Strengths and limitations

This is the first study that focused on long COVID patients with severe fatigue, one of the most common, disabling, and persistent symptoms in long COVID. Strengths of our study include the comprehensive assessment of both clinical and immune characteristics in long COVID patients with and without fatigue. This study is limited by the absence of a group of fully recovered acute COVID-19 patients, without signs of long COVID, and we, therefore, cannot confirm that our findings can solely be attributed to the disease condition of long COVID rather than being a recovery sign of acute COVID-19 3-6 months after hospitalization. It is encouraging that other studies have found that excessive signs of low grade inflammation and high CD8^+^ T-lymphocyte activity typify long COVID cases amongst the convalescent post COVID cases.^18, 20, 23^ Long COVID patients should be followed for a longer period of time evaluating both clinical and immunologic characteristics.

## 5. Conclusion

This study shows that long COVID patients with severe fatigue are most severely affected, with this group being characterized by many concurrent and persistent symptoms and by clear signs of immune alterations, specifically a definite state of low grade inflammation and high levels of CD8^+^ T-lymphocytes. Persistent symptoms and immune alterations were also present in long COVID patients without fatigue. Altogether the state of immune activation in long COVID shows resemblances to what has been earlier described in ME/CFS and MDD. The diversity of immune abnormalities indicates that personalized therapies combatting the diverse immune abnormalities may be required to alleviate the persisting disabling complaints of the patients.

## Data Availability

The dataset that support the findings of this study is not publicly available, but are available from the corresponding author upon reasonable request.

## Contributors

WAD and MEH shared senior authorship and contributed equally to this paper. All authors contributed to the acquisition, analysis, or interpretation of data. JCB performed the statistical analyses. AW performed the laboratory analysis. JCB, HAD, WAD, MEH drafted the manuscript. All authors critically revised and approved the final version of the manuscript. HAD, WAD, MEH provided supervision.

