## Supplemental Materials for "Severe fatigue as symptom of long COVID is characterized by increased expression of inflammatory genes in monocytes, increased serum pro-inflammatory cytokines, and increased CD8+ T-lymphocytes: A putative dysregulation of the immune-brain axis, the coagulation process, and auto-inflammation to expl"

| Table of Contents | page |
|---|---|
| Supplementary Methods | 1 |
| Corona symptom checklist | 1 |
| Supplementary Results | 2 |
| Unsupervised hierarchical cluster analysis using immune characteristics in long COVID patients | 2 |
| Table S1. Demographic and clinical characteristics | 3 |
| Table S2. Overview of the immune characteristics of each immunotype and the frequencies of the clinical variants in the immunotypes | 5 |
| Figure S1. Proportion of samples with a detectable value for a given cytokine and soluble cell surface molecule in the serum | 6 |
| Figure S2. Comparison of serum cytokine and soluble cell surface molecule levels across the three clinical variants of long COVID and healthy controls | 7 |
| Figure S3. Comparison of leukocyte and lymphocyte subsets across the three clinical variants of long COVID and healthy controls | 8 |
| Figure S4. Comparison of CD4+ T-lymphocyte subsets across the three clinical variants of long COVID and healthy controls | 9 |
| Figure S5. Unsupervised hierarchical cluster analysis using immune characteristics in long COVID patients (A) and the flow of patients within the clinical variants of long COVID over the six immunotypes of long COVID (B) | 10 |

### Supplementary Methods

#### Corona symptom checklist

We selected 12 typical long COVID symptoms from the corona symptom checklist, indicated in bold. *these symptoms were added to the questionnaire in a later stage and contain incomplete data.

*The following complaints may be experienced after SARS-CoV-2 infection. Check for each question whether you experienced the complaint at this moment and is new since the SARS-CoV-2 infection. When you experienced the complaints already before the infection and this did not change, please check "no". When the complaints worsened or is new after infection, please check "yes".*

| **Complaint:** | **Yes** | **No** |
|---|---|---|
| Do you experience vision problems? | | |
| **Do you experience dysgeusia?** | | |
| **Do you experience anosmia?** | | |
| **Do you experience excessive coughing?** | | |
| Do you experience phlegm? | | |
| Do you experience hoarseness? | | |
| **Do you experience dizziness or balance difficulties?** | | |
| Do you experience stool problems? | | |
| Do you experience miction problems? | | |
| Do you experience fatigue?* | | |
| Do you experience dyspnea?* | | |
| **Do you experience joint complaints?** | | |
| **Do you experience muscle weakness?** | | |
| **Do you experience reduced fitness?** | | |
| **Do you experience hair loss?** | | |
| Do you experience skin rash? | | |
| Do you experience claudication? | | |
| Do you experience sensory overload?* | | |
| **Do you experience concentration problems?** | | |
| **Do you experience memory problems?** | | |
| Do you experience headache?* | | |
| **Do you experience sleeping problems?** | | |
| Do you experience anxiety/nightmares? | | |
| **Do you experience tingling and or pain in extremities?** | | |
| Do you experience chest pain?* | | |

### Supplementary Results

#### Unsupervised hierarchical cluster analysis using immune characteristics in long COVID patients

We performed an unsupervised hierarchical cluster analysis to assess the patients on the basis of their immunotype. The most discriminative markers were added in the analysis and included the pro-inflammatory monocyte genes IL-6, CCL20, CXCL2, $CD8^+$ T-lymphocytes, and the percentage of naïve $CD4^+$ T-lymphocytes. Addition of the other found immune abnormalities did not discriminate the long COVID patients better. We were able to divide the patients into six different immunotypes (Figure S5A). The increase of pro-inflammatory cytokines was more or less equally divided over these immunotypes.

Figure S5B and Table S2 show that trends of a preferred link of the clinical variants of long COVID to specific immunotypes were visible. When we took all the immunotypes characterized by monocyte inflammatory gene activation together (immunotypes A, C, E, F), 25/35 (71.4%) of the severe long COVID patients fell into this group, while the immunotypes without monocyte inflammatory activation (immunotypes D,  B) only harbored 10/35 (29.6%) of the severe long COVID patients.

The immunotypes characterized by a naïve $CD4^+$ T-lymphocytopenia (types A, B, C) were linked predominantly with the moderately severe variant of long COVID and 15/24 (62.5%) of these patients fell into these immunotypes, while the immunotypes not characterized by a naïve $CD4^+$ T- lymphocytopenia (types D, E, F) only harbored 9/24 (38.5%) of the moderately severe long COVID patients.

The latter groups D, E, and F also had a predominance of the mild variant (8/10, 80.0%) and the highest prevalence of the mild long COVID patients (40.0%) was in group F, characterized by a monocyte inflammatory state and a $CD4^+$ *T-lymphocytosis*.

**Table S1. Demographic and clinical characteristics**

| | LC fatigue (n = 37) | LC non-fatigue | | |
|---|---|---|---|---|
| | | Moderately severe variant (n = 25) | Mild variant (n = 11) | P value |
| **Demographic characteristics** | | | | |
| Age (years) | 59.0 (56.0-66.5) | 60.0 (51.5-67.5) | 65.0 (59.0-68.0) | 0.5 |
| Male sex | 24 (64.9) | 18 (72.0) | 8 (72.7) | 0.8 |
| BMI (kg/m²) | 31.1 (27.0-34.2)<br>29.9 (26.4-32.8) | 28.4 (26.5-32.4)<br>27.3 (26.4-32.2) | 29.1 (26.0-34.5)<br>27.2 (26.3-32.4) | 0.5 |
| Comorbidities | | | | |
| ≥1 comorbidity | 32 (86.5) | 19 (76.0) | 10 (90.9) | 0.5 |
| Obesity (BMI≥30) | 16 (43.2) | 9 (36.0) | 4 (36.4) | |
| Diabetes | 9 (24.3) | 4 (16.0) | 2 (18.2) | |
| Cardiovascular disease and/or hypertension | 17 (45.9) | 9 (36.0) | 4 (36.4) | |
| Pulmonary disease | 9 (24.3) | 6 (24.0) | 2 (18.2) | |
| Renal disease | 9 (24.3) | 4 (16.0) | 1 (9.1) | |
| Gastrointestinal disease | 1 (2.7) | 1 (4.0) | 1 (9.1) | |
| Neurological disease | 4 (10.8) | 1 (4.0) | 5 (45.5) | |
| Malignancy | 4 (10.8) | 5 (20.0) | 3 (27.3) | |
| Autoimmune disease | 4 (10.8) | 4 (16.0) | 4 (36.4) | |
| Mental disorder | 1 (2.7) | 0 (0.0) | 0 (0.0) | |
| Ethnicity | | | | 0.7 |
| Caucasian | 28 (75.7) | 19 (79.2) | 10 (90.9) | |
| Other | 9 (24.3) | 6 (20.8) | 1 (9.1) | |
| Employed | 26 (70.3) | 15 (60.0) | 5 (45.5) | 0.5 |
| **Clinical characteristics at hospital admission** | | | | |
| Laboratory values | | | | |
| CRP (mg/L) | 80.0 (52.5-179.5) | 114.0 (45.0-196.0) | 78.0 (42.0-164.0) | 0.8 |
| Ferritin (ug/L) | 1196.0 (419.0-2205.5) | 948.0 (622.5-3600.8) | 977.0 (532.5-1610.5) | 0.7 |
| D-dimer (mg/L) | 1.1 (0.747-1.9) | 0.98 (0.61-2.4) | 0.64 (0.24-1.1) | 0.3 |
| Lymphocytes absolute count ($10^9$/L) | 0.96 (0.80-1.2) | 0.70 (0.57-1.0) | 0.86 (0.48-0.94) | 0.1 |
| Treatment | | | | - |
| None | 1 (2.7) | 0 (0.0) | 0 (0.0) | |
| Steroids | 36 (97.3) | 25 (100.0) | 11 (100.0) | |
| Anti-inflammatory | 12 (32.4) | 10 (40.0) | 3 (27.3) | |
| Antivirals | 1 (2.7) | 1 (4.0) | 0 (0.0) | |
| Covalescent plasma | 1 (2.7) | 1 (4.0) | 0 (0.0) | |

| | | | | |
|---|---|---|---|---|
| Hydroxy)chloroquine | 0 (0.0) | 0 (0.0) | 0 (0.0) | |
| Oxygen supplementation | 37 (100.0) | 25 (100.0) | 11 (100.0) | - |
| HFNC | 18 (48.6) | 14 (56.0) | 3 (27.3) | 0.4 |
| IMV | 16 (43.2) | 15 (60.0) | 4 (36.4) | 0.3 |
| ICU admission | 18 (48.6) | 17 (68.0) | 5 (45.5) | 0.3 |
| LOS ICU (days | 11.0 (9.0-16.0) | 14.0 (9.0-20.0) | 9.0 (4.0-21.0) | 0.5 |
| LOS hospital (days) | 17.0 (9.0-26.0) | 20.0 (10.5-29.5) | 13.0 (9.0-14.0) | 0.3 |
| **Collection of blood samples** | | | | |
| Time since hospital discharge (days) | 106.0 (92.5-118.5) | 114.0 (92.5-168.5) | 106.0 (86.0-130.0) | 0.6 |
| Vaccinated against COVID-19 | 26 (78.8) | 19 (86.4) | 6 (66.7) | 0.5 |

Data are presented as median (interquartile range) or number (percentage). Group comparisons were performed using a Kruskal-Wallis test and Fisher-Freeman-Halton exact test, as appropriate. LC fatigue: patients experienced fatigue at the time of collecting blood samples. LC non-fatigue, moderately severe variant: patients experienced impaired fitness without fatigue. LC non-fatigue, mild variant: patients without impaired fitness and fatigue but reporting other symptoms. CRP: C-reactive protein, HFNC: high flow nasal cannula, ICU: intensive care unit, IMV: invasive mechanical ventilation, LC: Long COVID, LOS: length of stay.

**Table S2. Overview of the immune characteristics of each immunotype and the frequencies of the clinical variants in the immunotypes**

| Immunotype | Monocyte activation | $CD8^+$ T-lymphocytes | CD45RO- naïve $CD4^+$ T-lymphocytes | Description | LC clinical variant |
|---|---|---|---|---|---|
| **A (n=11)** | + | + | -- | $CD4^+$ lymphocytopenia ++<br>Monocyte inflammation+<br>$CD8^+$ T-lymphocytes + | 5/35 (14.3%) of LC fatigue variant<br>5/24 (20.8%) of LC non-fatigue, moderately severe variant<br>1/10 (10.0%) of LC non-fatigue, mild variant |
| **B (n=11)** | normal | + | -- | $CD4^+$ lymphocytopenia ++<br>$CD8^+$ T-lymphocytes + | 4/35 (11.4%) of LC fatigue variant<br>7/24 (29.2%) of non-fatigue, moderately severe variant<br>0/10 (0%) of non-fatigue, mild variant |
| **C (n=8)** | + | normal | -- | Monocyte inflammation+<br>$CD4^+$ lymphocytopenia ++ | 4/35 (11.4%) of LC fatigue variant<br>3/24 (12.5%) of LC non-fatigue, moderately severe variant<br>1/10 (10.0%) of LC non-fatigue, mild variant |
| **D (n=10)** | normal | normal | normal | Normal | 6/35 (17.1%) of LC fatigue variant<br>2/24 (8.3%) of LC non-fatigue, moderately severe variant<br>2/10 (20.0%) of LC non-fatigue, mild variant |
| **E (n=13)** | ++ | + | normal | Monocyte inflammation ++<br>$CD8^+$ T-lymphocytes + | 10/35 (28.6%) of LC fatigue variant<br>1/24 (4.2%) of LC non-fatigue, moderately severe variant<br>2/10 (20.0%) of LC non-fatigue, mild variant |
| **F (n=16)** | + | normal | **++** | $CD4^+$ *T-lymphocytosis*<br>Monocyte inflammation + | 6/35 (17.1%) of LC fatigue variant<br>6/24 (25.0%) of LC non-fatigue, moderately severe variant<br>4/10 (40.0%) of LC non-fatigue, mild variant |

In the table + and ++ (upregulation) indicate an upregulation of the characteristic compared to the healthy controls; - and -- (downregulation) indicate a downregulation of the characteristic compared to the healthy controls. LC: long COVID.

**Figure S1. Proportion of samples with a detectable value for a given cytokine and soluble cell surface molecule in the serum**

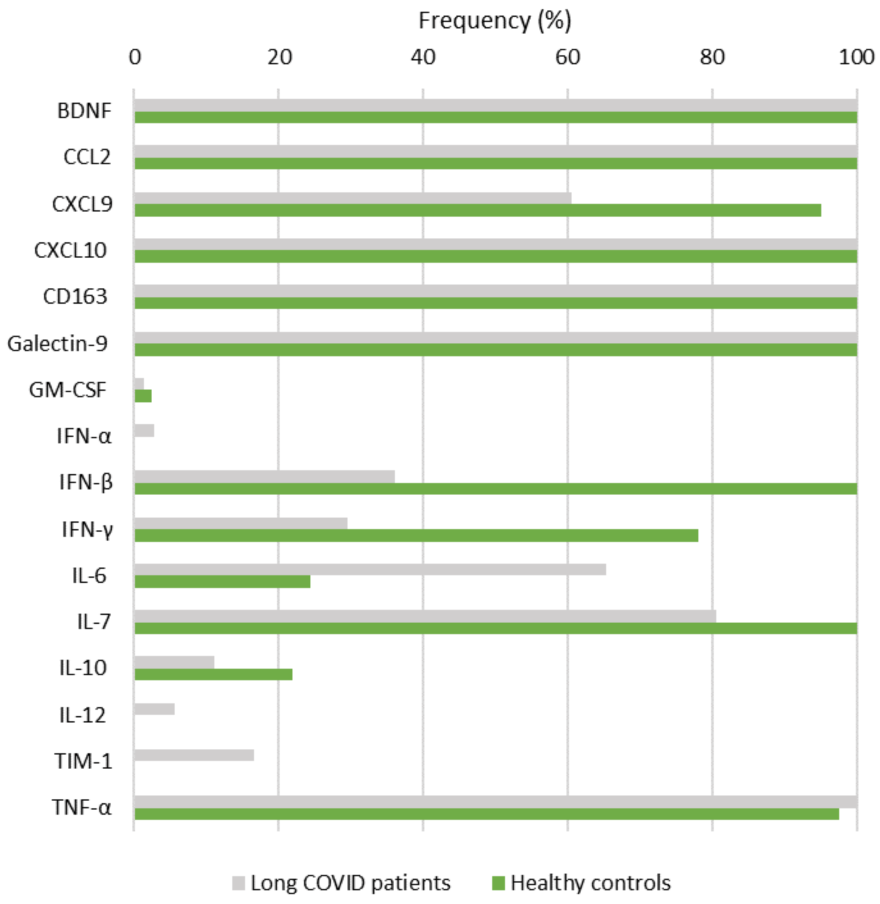


BDNF: brain-derived neurotrophic factor, CCL: C-C motif chemokine ligand, CXCL: C-X-C motif chemokine ligand, CD163: cluster of differentiation, IL: interleukin, GM-CSF: granulocyte macrophage-colony stimulating factor, IFN: interferon, TIM: T-cell immunoglobulin and mucin domain 1 (TIM-1), TNF-α: tumor necrosis factor-alpha.

**Figure S2. Comparison of serum cytokine and soluble cell surface molecule levels across the three clinical variants of long COVID and healthy controls**

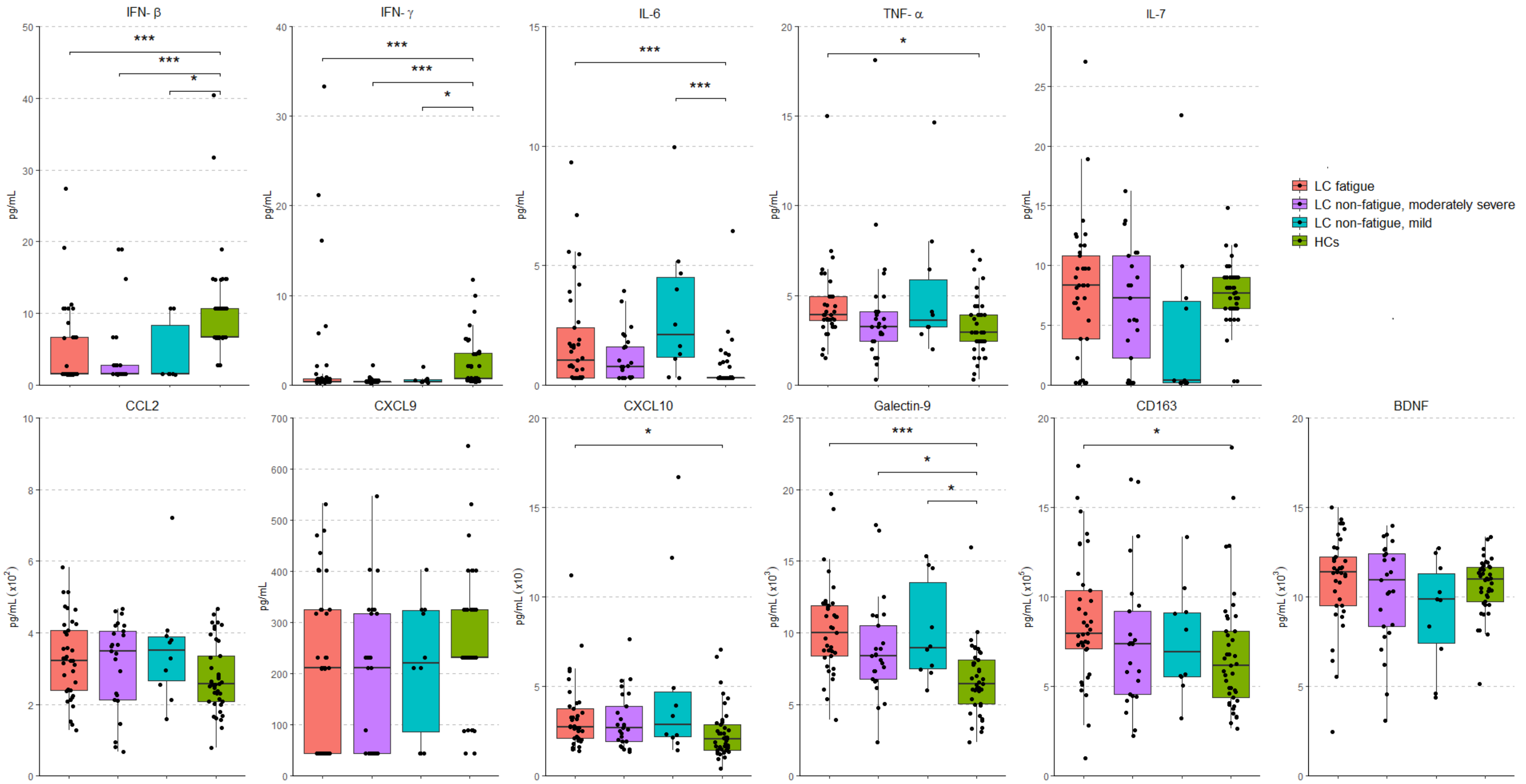


Group comparisons were performed using the Kruskal-Wallis test for CCL2 (overall p-value=0.19), CXCL9 (p=0.04) CXCL10 (p=0.01), IL-6 (p<0.001), IL-7 (p=0.2), TNF-α (p=0.02), IFN-β (p<0.001), IFN-γ (p<0.001), Galectin-9 (p<0.001), CD163 (p=0.04), and BDNF (p=0.3). Post-hoc analysis was performed using Bonferroni correction for multiple comparisons, a significant group difference is denoted by *p<0.05, **p<0.01, ***p<0.001. LC fatigue: patients experienced fatigue at the time of collecting blood samples. LC non-fatigue, moderately severe variant: patients experienced impaired fitness without fatigue. LC non-fatigue, mild variant: patients without impaired fitness and fatigue but reporting other symptoms. HCs: Healthy controls, LC: Long COVID. BDNF: brain-derived neurotrophic factor, CCL: C-C motif chemokine ligand, CXCL: C-X-C motif chemokine ligand, CD163: cluster of differentiation, IL: interleukin, GM-CSF: granulocyte macrophage-colony stimulating factor, IFN: interferon, TIM: T-cell immunoglobulin and mucin domain 1 (TIM-1), TNF-α: tumor necrosis factor-alpha.

**Figure S3. Comparison of leukocyte and lymphocyte across the three clinical variants of long COVID and healthy controls**

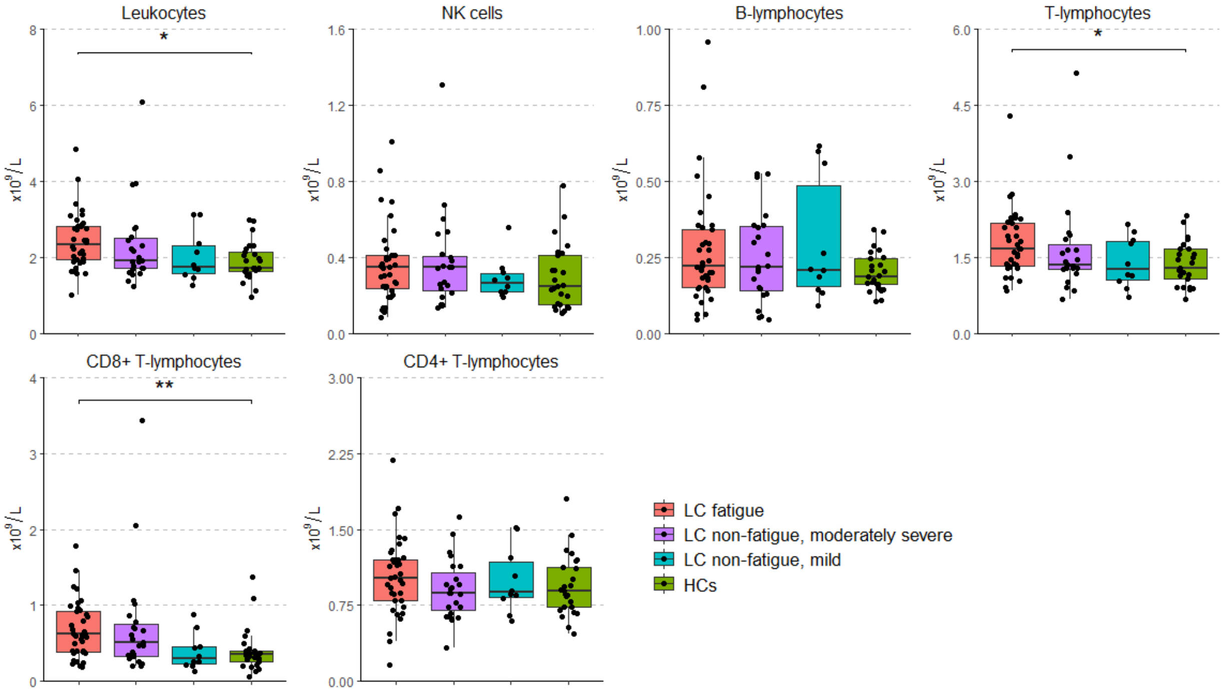


Group comparisons were performed using the Kruskal-Wallis test for the counts of leukocytes ($CD45^+$) (overall p-value= 0.02), NK ($CD3^-CD16^+CD56^+$) cells (p=0.4), B-lymphocytes ($CD19^+$) (p=0.6), T-lymphocytes ($CD3^+$) (0.04), $CD8^+$ T-lymphocytes ($CD3^+CD8^+$) (p=0.002), and $CD4^+$ T-lymphocytes ($CD3^+CD4^+$) (p=0.5). Post-hoc analysis was performed using Bonferroni correction for multiple comparisons, a significant group difference is denoted by *p<0.05, **p<0.01, ***p<0.001. LC fatigue: patients experienced fatigue at the time of collecting blood samples. LC non-fatigue, moderately severe variant: patients experienced impaired fitness without fatigue. LC non-fatigue, mild variant: patients without impaired fitness and fatigue but reporting other symptoms. HCs: Healthy controls, LC: Long COVID. NK cells: natural killer cells.

### Figure S4. Comparison of CD4$^+$ T-lymphocyte subsets across the three clinical variants of long COVID and healthy controls

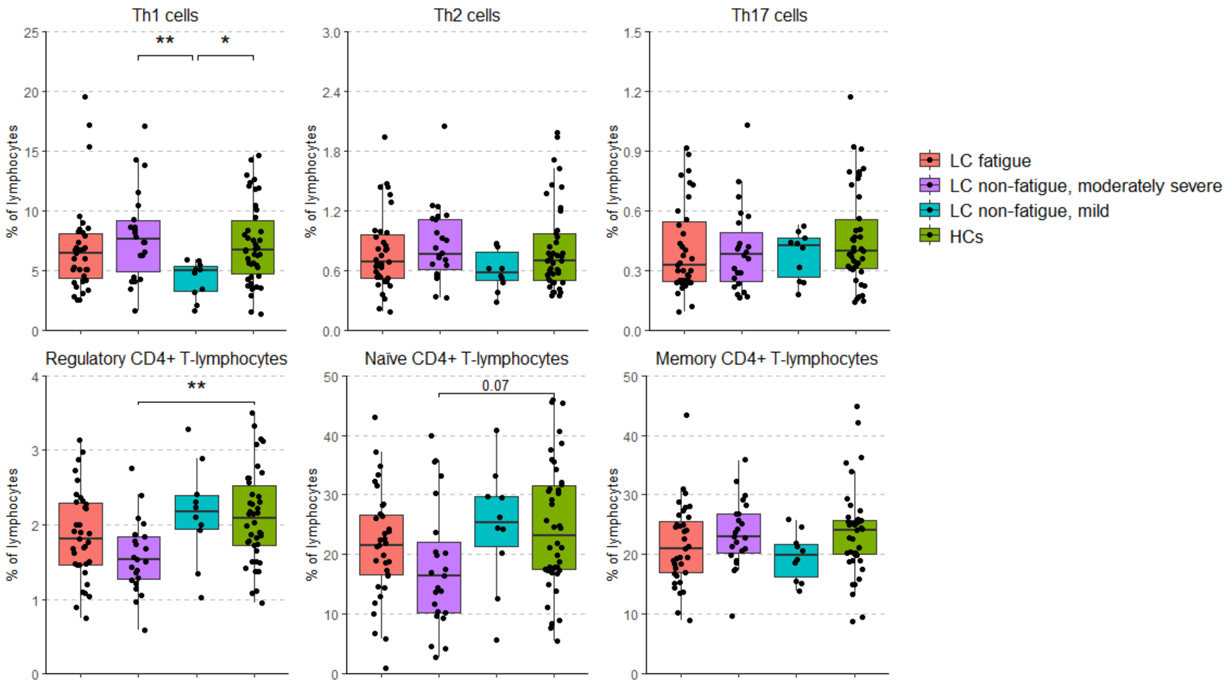


Group comparisons were performed using the Kruskal-Wallis test for the percentage of Th1 (CD3$^+$CD4$^+$IFNγ$^+$) cells (overall p-value=0.01), Th2 (CD3$^+$CD4$^+$IL4$^+$) cells (p=0.3), Th17 (CD3$^+$CD4$^+$IL17A$^+$) cells (p=0.6), Treg (CD3$^+$CD4$^+$CD25hiFOXP3$^+$) cells (p=0.006), naïve (CD3$^+$CD4$^+$CD45RO$^-$) CD4+ T-lymphocytes (p=0.07), and memory (CD3$^+$CD4$^+$CD45RO$^+$) CD4+ T-lymphocytes (p=0.2), relatively to the total lymphocytes. Post-hoc analysis was performed using Bonferroni correction for multiple comparisons, significant group difference is denoted by *p<0.05, **p<0.01, ***p<0.001. LC fatigue: patients experienced fatigue at the time of collecting blood samples. LC non-fatigue, moderately severe variant: patients experienced impaired fitness without fatigue. LC non-fatigue, mild variant: patients without impaired fitness and fatigue but reporting other symptoms. HCs: Healthy controls, LC: Long COVID.

**Figure S5. Unsupervised hierarchical cluster analysis using immune characteristics in long COVID patients (A) and the flow of patients within the clinical variants of long COVID over the six immunotypes of long COVID (B)**

**A**

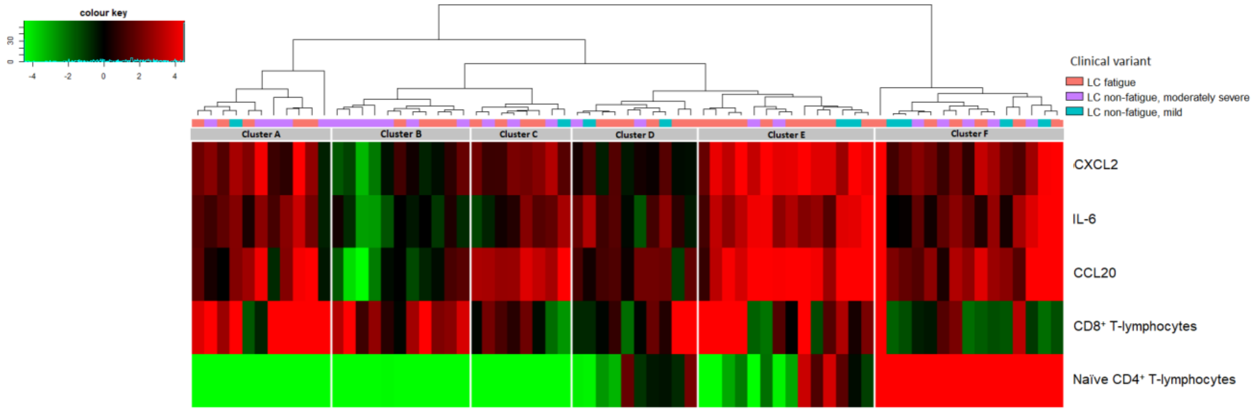


**B**

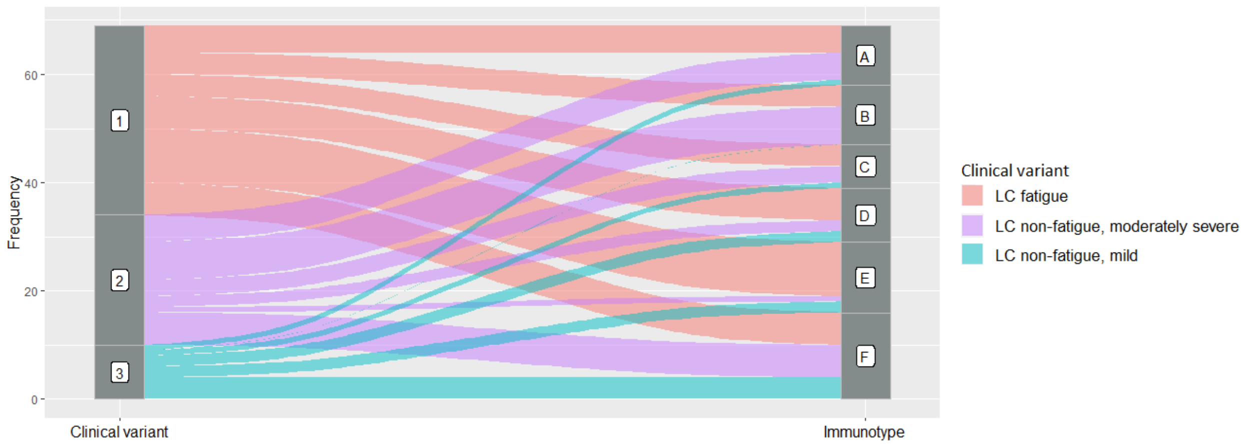


S5A: The most discriminative immune markers were entered in the analysis and included the pro-inflammatory monocyte genes IL-6, CCL20, and CXCL2, the number of $CD8^+$ T-lymphocytes, and the percentage of naïve $CD4^+$ T-lymphocytes ($CD45RO^-$). The analysis revealed six immunotypes, representing clusters of patients with similar immune expressions. S5B: The flow of patients within clinical variants of long COVID over the six immunotypes. LC fatigue: patients experienced fatigue at the time of collecting blood samples. LC non-fatigue, moderately severe variant: patients experienced impaired fitness without fatigue. LC non-fatigue, mild variant: patients without impaired fitness and fatigue but reporting other symptoms. LC: Long COVID.
